# Modelling the impact of a school-based influenza vaccination programme under uncertainty in seasonal transmission dynamics

**DOI:** 10.64898/2026.11.22.26368237

**Authors:** Eliran Boksenbojm, Oliver Eales, Allen C Cheng, James McCaw, Freya Shearer

**Author notes:** These authors contributed equally to this work (Eliran Boksenbojm, Oliver Eales). Corresponding authors: Freya Shearer, Eliran Boksenbojm.

## Abstract

School-aged children are thought to be an important driver of influenza transmission. In this paper we study the direct and indirect health effects of a school-based live-attenuated influenza vaccination (LAIV) programme in Australia, using a stochastic, age-stratified model of seasonal influenza transmission. The model was calibrated using Approximate Bayesian Computation (ABC) in order to capture uncertainty in influenza infection dynamics. The calibrated model was used to evaluate programmes varying in target age group, vaccination coverage and vaccine effectiveness. On average, school-based LAIV programmes reduced infections and hospitalisations at the population level across all scenarios, with greater reductions at higher coverage and vaccine effectiveness and when both primary- and secondary-school-aged children were targeted. However, estimated effects varied substantially across plausible transmission dynamics, particularly indirect effects — for some programme scenarios, we estimated that in non-target age groups both near complete suppression or a small increase in infections were possible outcomes. For all scenarios, we estimated that school-based LAIV programmes would lead to a reduction in infections and hospitalisations in target age groups.

## 1 Introduction

Influenza viruses circulate continuously through the human population with transmission driven by immunological, demographic, evolutionary, and seasonal processes. In temperate regions, where seasonal processes are stronger compared to the tropics, influenza viruses typically cause yearly epidemics with activity peaking during the colder months. In tropical and subtropical regions, seasonality is less distinct and various transmission regimes are observed including year-round activity or multiple peaks [1]. While the majority of influenza infections result in mild illness [2], a small proportion of infections result in more severe outcomes (e.g. hospitalisation and death); due to the high rates of influenza infections annually, this small proportion leads to substantial mortality and morbidity [3, 4]. While vaccination against influenza can be effective, adaptive evolution of influenza viruses leads to new antigenically distinct strains (“antigenic drift”) that can reinfect previously immune individuals [5]. As such, influenza vaccines require frequent updates (vaccine composition assessed twice a year for the northern and southern hemispheres influenza seasons) [6], and many countries including Australia recommend annual vaccination for all individuals (except very young children) [7]. The effectiveness of seasonal influenza vaccines at preventing infection and hospitalisation varies year-on-year depending on how well the vaccine strain composition matches with circulating strains, among other factors [8]. For standard injectable inactivated influenza vaccines (IIV), effectiveness is generally measured to be around 40–60% against medically-attended illness and 50–70% against hospitalisation [9].

The clinical severity of influenza infections is heterogeneous with age, with higher rates of severe outcomes observed in older individuals [10]. Thus while vaccination is recommended to all age groups, vaccination campaigns often prioritise groups most at risk of severe infection, including older individuals (e.g. by offering free and subsidised vaccines); in Australia vaccination coverage in 2025 was highest in individuals aged 65 and over (60.7%) and lowest in ages 12–*<*18 (12.1%) [11]. While older age groups are at greater risk of severe outcomes, it is thought that transmission dynamics are predominantly driven by younger individuals, particularly school-aged children, who have higher rates of social contact [12, 13]. Increasing influenza vaccination coverage in school-aged children is therefore considered to have substantial potential to reduce transmission, indirectly leading to reductions in severe outcomes in more at-risk age groups (and unvaccinated school children). The advent of live attenuated influenza vaccines (LAIV) over the past decade, has led to an interest in paediatric vaccination strategies, particularly as intranasal delivery of LAIV may be more acceptable to children, and when combined with a school-based rollout programme, high coverage could be achieved in this age group. Furthermore, recent vaccine effectiveness (VE) studies of LAIV (e.g. [14]) indicate that VE is commensurate, if not higher, than standard IIV. A 2024 study from the United Kingdom reported vaccine effectiveness against primary-care attended influenza infection of 66% (95% CI: 53%–76%) in children aged 2–17 years (predominately receiving LAIV) and 47% (95% CI: 37%–56%) in adults aged 18-64 (predominantly receiving IIV) [14].

Assessing the likely effect of school-based LAIV programmes, including a quantification of uncertainty, is required to inform policy decisions. However, it is non-trivial to estimate the potential impact of such a programme due to the non-linear processes that drive influenza transmission and limitations in epidemiological data. For example, information on the true burden of influenza infections is scarce, as data is collected predominantly on downstream outcomes representing only a fraction of overall infections (e.g. cases, hospitalisations, deaths). Additionally, there are likely high levels of heterogeneity in the level of pre-existing immunity to influenza strains circulating in a given season — serological data that would allow this heterogeneity to be quantified are not routinely collected. Further, this pre-existing immunity to circulating strains can decay over time due to either: the waning of short-term immunity [15]; or the emergence of novel strains via antigenic drift. Considering the uncertainty in these dynamics is paramount for robustly quantifying uncertainty in the effectiveness of school-based vaccination programmes.

Here we develop a stochastic model of seasonal influenza transmission stratified by age, vaccination, and prior immunity status. The model is calibrated using a two-step process. First, we use Approximate Bayesian Computation (ABC) with a set of summary statistics based on the expected influenza infection dynamics in the Australian context, allowing a wide range of uncertainty in infection dynamics to be considered. Then, for each infection trajectory accepted by the ABC algorithm, we computed age-stratified infection hospitalisation rates according to observed rates of hospitalisation by age. The model was then used to estimate the population-level and age-specific impact of school-based LAIV programmes, and examine how results vary under different assumptions regarding vaccine effectiveness, vaccine coverage and target age groups. This work was conducted as part of a multi-model exercise coordinated by the Australia–Aotearoa Scenario Modelling Hub.

## 2 Results

### 2.1 Baseline infection dynamics of influenza

Approximate Bayesian Computation (ABC) identified a set of parameter combinations (model particles) that reproduced key characteristics of seasonal influenza epidemics in Australia under the current IIV vaccination programme—the Baseline scenario (Figure 1). Daily infection dynamics of accepted simulations over the one-year horizon were consistent with epidemic dynamics observed in influenza case notification data from Australian temperate regions (Figure S8). Specifically, 518 of 70,000 model particles met our ABC acceptance criteria (see Methods). These criteria targeted Baseline epidemic trajectories with a peak timing between day 147 (May 27) and day 289 (October 16) (Figure 1, panel **A**), an annual cumulative infection proportion between 10–45% (Figure 1, panel **B**), and IIV VE against infection between 38–50% (Figure 1, panel **C**). An example of the daily and cumulative infection incidence for such an accepted particle is shown in Figure S24 and Figure S25, respectively.

**Figure 1:**
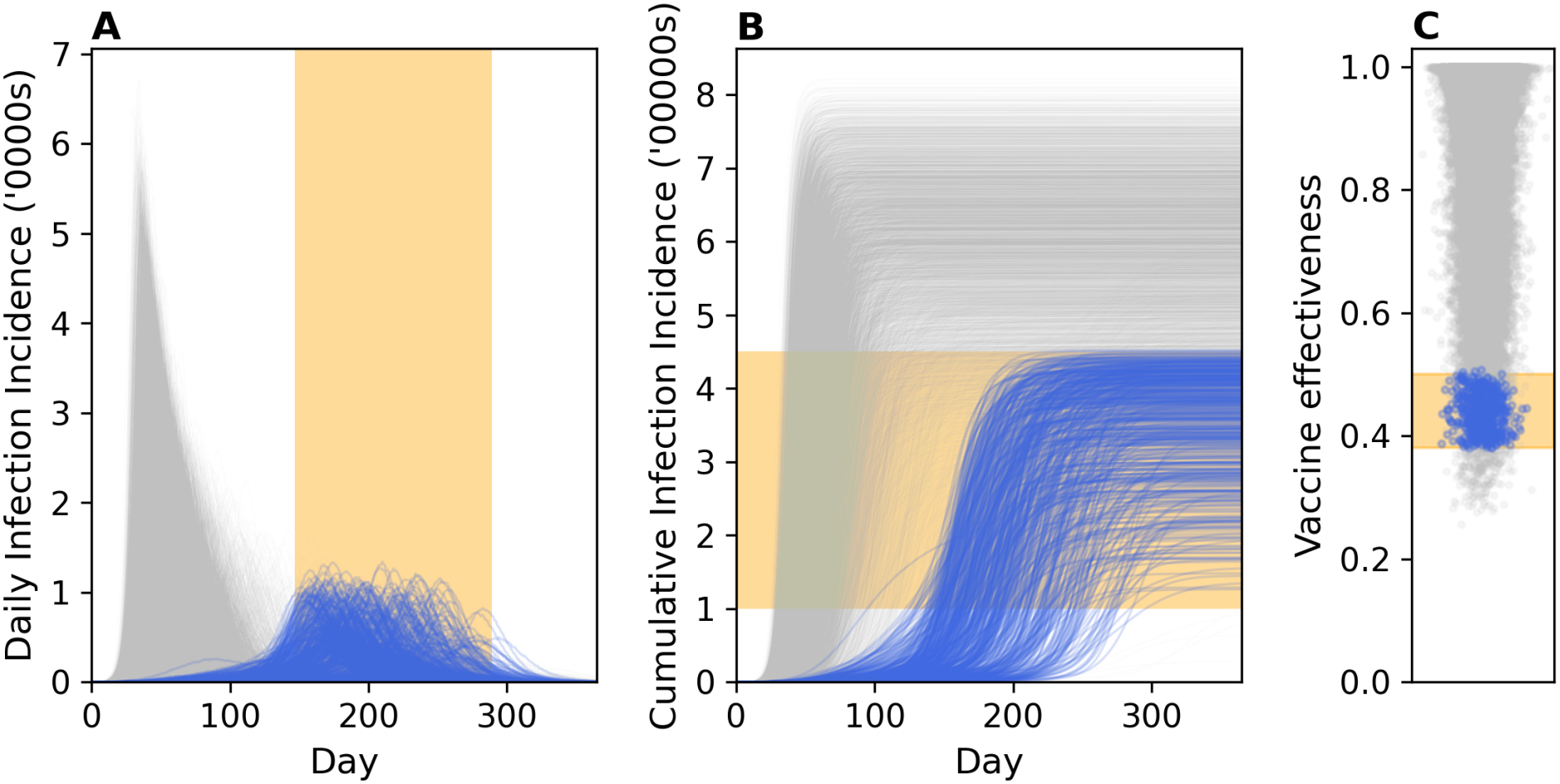
Model outputs from randomly selected model particles sampled from the Approximate Bayesian Computation (ABC) priors using Latin Hypercube Sampling. Panel **A** shows the daily infection incidence in tens of thousands, panel **B** the cumulative infection incidence in hundreds of thousands, and panel **C** the IIV VE against infection. Blue lines (dots) represent outputs from accepted model particles and grey lines (dots) rejected ones. Orange boxes show the calibration targets used for ABC acceptance.

The relationship between accepted parameter combinations were highly complex due to the high dimensionality of the system (eight free parameters), but we investigated their pairwise distributions (Figure S7). We observed moderate correlation between the fraction with pre-existing immunity (*S*^1^ compartment in Methods) and the average annual transmissibility (*β*_0_), and moderate negative correlation between the fraction with pre-existing immunity and the degree of seasonality (*β*_1_). Accepted parameter values of *β*_0_ corresponded to average annual values (averaged over seasonal variation) of the basic reproduction number — *R*_0_, the expected number of secondary infections caused by an initial infection in a fully susceptible population — between 1.58 and 2.34, with a median value of 1.74. Accepted parameter combinations of *β*_0_ and *β*_1_ corresponded to maximum values (i.e. when transmissibility is highest during the season) of *R*_0_ between 1.67 and 3.00, with a median of 2.11 (see Figure S29).

As expected, due to their higher rates of assumed contacts and lower Baseline vaccination coverage, the highest infection rates were in school-aged children with annual infection attack rates per 100,000 ranging from 11,162–63,986 for children aged 5–*<*12 and 14,096–70,570 for ages 12–*<*18 (Figure S27). The lowest infection rates were in the oldest age-groups with infection attack rates per 100,000 ranging from 2,400–23,424 for ages 65–*<*80 and 1,616–16,283 for ages 80+. The greatest number of total infections was in ages 18–*<*65 reflecting the larger population size of this age group (see Figure S26). Trends in infections in school-aged children led the overall dynamics of the epidemics with peaks in infections in school-aged children leading peaks in other age groups by less than 1 week (Figures S30, S31, and S28).

### 2.2 Population-level impacts of school-based vaccination programmes on influenza infection dynamics

For all LAIV programme scenarios examined (see Section 4.5 for details), we estimated median reductions in infections at the population-level compared to the Baseline scenario. However, we estimated substantial variability around those estimates, including an increase in infections for a subset of model particles (Figure 2). For each of the (518) accepted particles, we simulated influenza infection dynamics for each scenario (50 simulations each) and compared the overall infection burden to simulations of the Baseline scenario (50 simulations as well). Outcomes were summarised by their median over 50 simulations to reduce uncertainty in the estimated impact of LAIV programmes due to stochastic (process) noise, and thus better characterise the uncertainty due to parameter uncertainty. Minimal variance was observed across 50 simulations and hence deemed sufficient for reducing stochastic noise (Figures S1–S4). The daily infection incidence as well as the infection attack rate across all simulated scenarios can be found in the the supplementary material (Figures S22 and S13).

**Figure 2:**
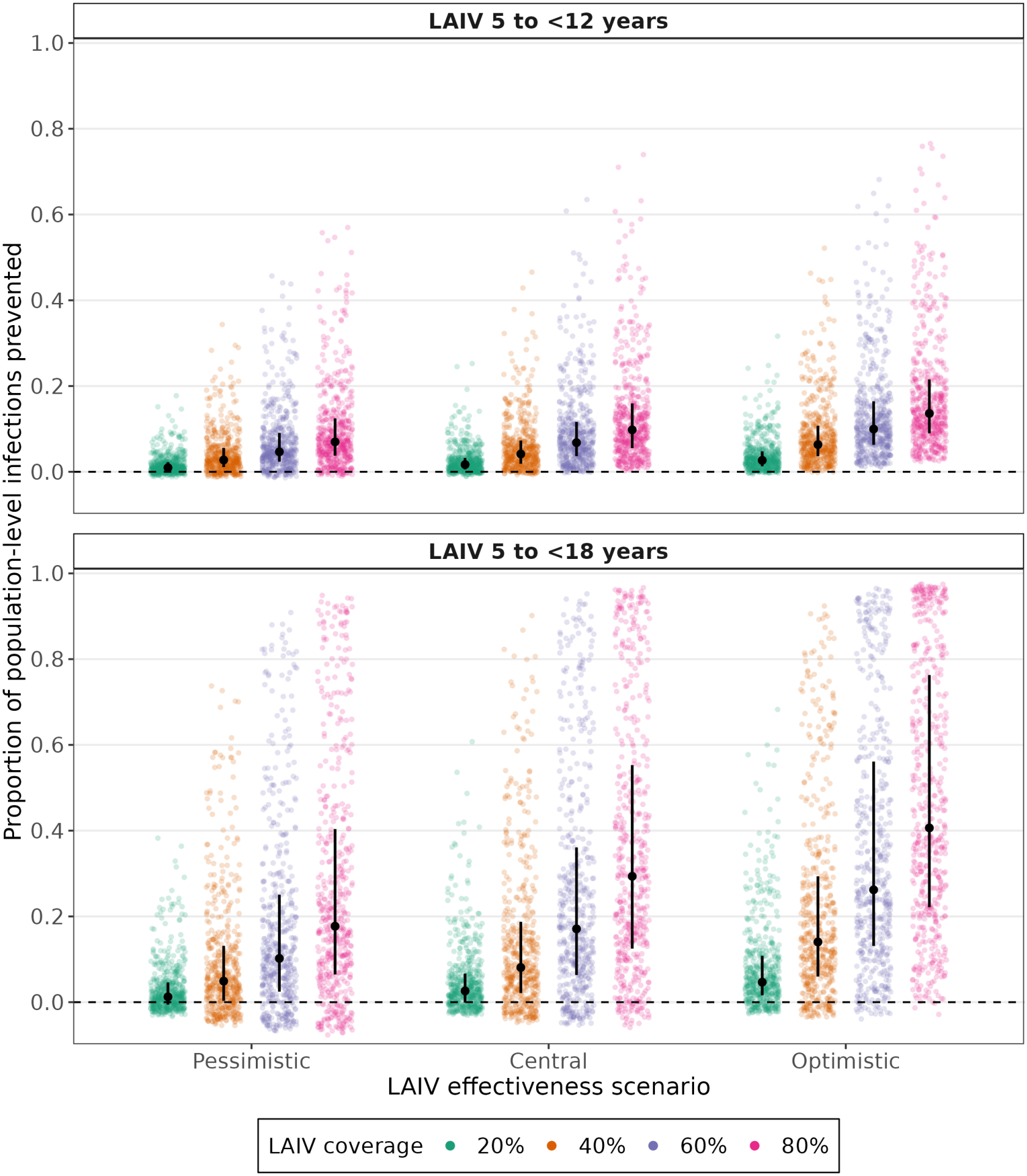
Proportion of population-level influenza infections prevented by school-based LAIV vaccination programmes under different assumptions about LAIV effectiveness and coverage. Results are shown for LAIV administered to children aged 5–*<*12 years (top) and 5–*<*18 years (bottom), under Pessimistic, Central, and Optimistic assumptions about LAIV effectiveness and coverage levels of 20%, 40%, 60%, and 80%. The proportion of infections prevented was calculated relative to the corresponding Baseline simulation. Coloured points show results for individual model particles, taking the median across replicate runs. Black points show the median across model particles, with vertical lines indicating the interquartile range.

As expected, the magnitude of median reductions in overall infections depended strongly on the LAIV coverage, LAIV effectiveness against infection, and target age group(s). Across all LAIV scenarios, vaccinating both primary and secondary school children (ages 5–*<*18) prevented a larger median proportion of infections overall than vaccinating only primary school children (ages 5–*<*12), by a factor ranging from 1.30 to 3.00 (Figure 2). For example, 4.2% of infections were prevented in a scenario of Central LAIV effectiveness (44–69%) and 40% coverage when LAIV was rolled out to primary school children only, which increased to 8.1% for a scenario where LAIV was also rolled out to secondary school children. Expected benefits increased with increasing LAIV coverage and effectiveness. For an LAIV programme targeting primary school children only and Central LAIV effectiveness, the median proportion of infections prevented increased from 1.7% to 9.8% as coverage increased from 20% to 80%. The greatest overall effects were observed under the most Optimistic vaccine assumptions (coverage 80%, LAIV effectiveness 63–88%) with median reductions of 13.6% (primary only) and 40.6% (both primary and secondary). In a sensitivity analysis we found that the effect of school-based LAIV programmes was highly dependent on the Baseline infection attack rate. An LAIV programme in primary and secondary school children (with Central VE and 60% coverage) led to near complete suppression of the epidemic for a high proportion of particles when the Baseline attack rate was lower (10–25%); whereas when the Baseline attack rate was higher (35–45%) there was a far greater proportion of particles in which the number of infections in non-target age groups increased (Figure S19). In additional sensitivity analyses, we found that school-based LAIV programmes led to greater reductions in infections (on average) when: seasonality in transmission was weaker (Figure S15); mean transmissibility (*R*_0_) was lower (Figure S16); maximum transmissibility was lower (Figure S17); maximum transmissibility occurred earlier in the season (Figure S18); the Baseline peak infection incidence time was later (Figure S21); and waning of pre-existing immunity was slower (Figure S20).

We estimated a high degree of uncertainty in infections prevented across all LAIV scenarios. This uncertainty increased for scenarios with higher levels of LAIV coverage and effectiveness or broader coverage (i.e. targeting both primary and secondary school children). For instance, the 95% interval width of 24.4% (−0.2%–24.2% infections prevented) when only primary school children were targeted, increased to 71.7%(−3.6%–68.1% infections prevented) when secondary school children were also targeted (Central LAIV effectiveness and 40% coverage). While the 95% credible intervals were substantially overlapping across LAIV scenarios (due to the high degree of variability in infections prevented across model particles within each scenario), when outputs were examined by scenario and model particle, the ranked impact of each particle was similar across scenarios (Figures S13 and S5). That is, the same particles generally led to the greatest (least) proportion of infections prevented across all scenarios. This provides us with strong evidence that the ranking of scenarios in terms of expected impact is robust.

The impact of school-based LAIV programmes was greatest in the target age groups, resulting from a combination of both direct and indirect effects (Figure 3A). Infections were also prevented in non-target age groups (such as older adults) in all scenarios, for the majority of model particles, indicating indirect effects. For instance, in the LAIV scenario of Central VE (44–69%) and 40% coverage targeting children aged 5–*<*18, we estimated a median reduction in infections of 26.8% for those aged 12–*<*18, and 3.0% in ages 65–*<*80. However, for a fraction of model particles, an increase in infections was observed in non-target age groups for LAIV scenarios compared to Baseline, indicating potentially negative impacts of LAIV programme introduction. As this effect was computed from the median over 50 stochastic simulations for each model particle, it is not simply due to process noise. For a programme targeting children aged 5–*<*18, with Central VE and 20% coverage, the median proportion of infections prevented was approximately zero for all non-target age groups. However, an increase in infections (relative to Baseline) was estimated in (for example) those aged 65–79 years of greater than 5.1% for 10% of model particles. As LAIV coverage increased, the median proportion of infections prevented in non-target age groups increased—with some model particles leading to near complete suppression of transmission—but so too did the potential negative impacts; at 80% coverage we estimated an increase in infections in ages 65–79 of greater than 13.3% for 10% of model particles. These age-specific effects translated to a portion of model particles producing negative impacts at the population-level (ranging from 25.9% for 20% coverage to 8.1% for 80% coverage). The population-level infections and hospitalisations reductions across all modelled scenarios are shown in Table S1 and Table S2, respectively.

**Figure 3:**
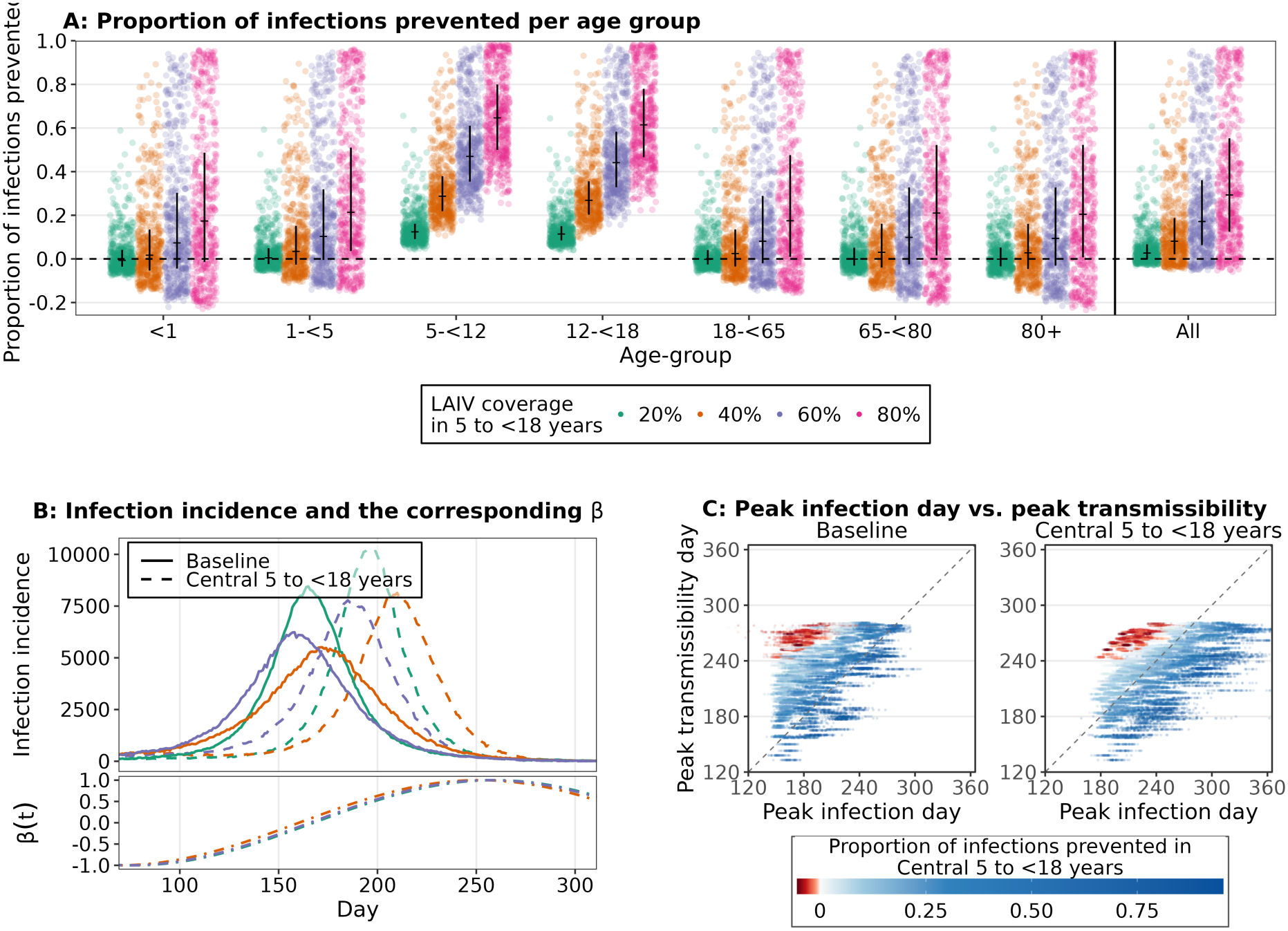
Age-stratified overview of infections prevented, with additional analysis of the results exhibiting increases in infections. (**A**) Age-stratified proportion of infections prevents for the scenarios with Central LAIV effectiveness against infection, with LAIV administered to ages 5–*<*18, with the corresponding coverage shown in different colours. (**B**) The transmissibility *β*(*t*) (bottom) for three particles giving rise to an increase in the infected population proportion over the course of the epidemic. The corresponding daily infection incidence for those three particles is shown (top) for the Baseline scenario as well as the Central LAIV effectiveness, 60% LAIV coverage in ages 5–*<*18. (**C**) Day of peak of transmissibility (i.e. peak of *β*(*t*)) versus day of peak infection incidence. Each of the 518 accepted particles is represented by 50 points, corresponding to its 50 simulation runs. Colour indicates the proportion of infections prevented in the 60% LAIV coverage, 5–*<*18, Central LAIV effectiveness scenario relative to Baseline. Particles associated with in increase in infections are clustered in the top left, corresponding to an early peak in infection incidence and late peak in transmissibility.

The underlying mechanism for the negative impacts was due to interactions between the timings of realised epidemics and seasonal forcing (that scales relative transmissibility). In the Baseline scenario, the model considers a wide range of parameter combinations that give rise to typical influenza epidemics in the Australian setting, including the strength and timing of seasonal forcing over the year. The model particles that led to negative impacts in non-target age groups for LAIV scenarios all exhibited similar dynamics in their corresponding Baseline scenarios: the peak in modelled infections occurred earlier in the season than the peak in seasonal forcing (See Figure 3 **B** and **C**). LAIV scenarios led to reduced transmission early in the year/season (due to higher vaccination coverage overall), thus shifting the epidemic to occur later in the season— into a period in which the relative transmissibility was higher, resulting in more infections in non-target age groups than at Baseline. In contrast, model particles that led to near complete epidemic suppression in LAIV scenarios exhibited Baseline peaks in modelled infections around the same time as their peak in relative transmissibility. For these particles, LAIV scenarios also shifted the epidemics to occur later, into a period in which relative transmissibility was lower (Figure S33). In sensitivity analyses, we found less uncertainty in our estimates (of the impact of school-based LAIV programmes) when seasonal changes in transmissibility were less gradual (i.e. more rapid change in transmissibility at season onset). However, some parameter combinations still led to near complete suppression or increases in infections in non-target age groups (relative to Baseline) for all patterns of seasonality in transmission considered (Figure S32).

### 2.3 Impact of the timing of school-based LAIV rollout

While our main analysis assumed an early rollout of vaccines to school-age children during term 1 (days 59–100, late-February to early-April), well ahead of the expected influenza season onset, we also considered a later rollout in term 2 (days 110–170, mid-April to mid-June). The median impact of an LAIV programme was similar for both rollout timings (Figure 4). The more striking result of this analysis was the reduction in uncertainty in the estimated proportion of infections prevented for a term 2 rollout compared to term 1. The timing of the term 2 rollout was later in the year, during a period in which, for some model particles, infections were already increasing (Figure 4); as such the LAIV programme had less impact on early transmission, and so epidemics were not shifted as late in the season (Figure S12). Consequently, fewer model particles produced epidemics under LAIV scenarios that: (1) resulted in increased infections in non-target age groups; or (2) greatly suppressed infections overall. This substantially reduced the range of possible outcomes and hence reduced uncertainty overall.

**Figure 4:**
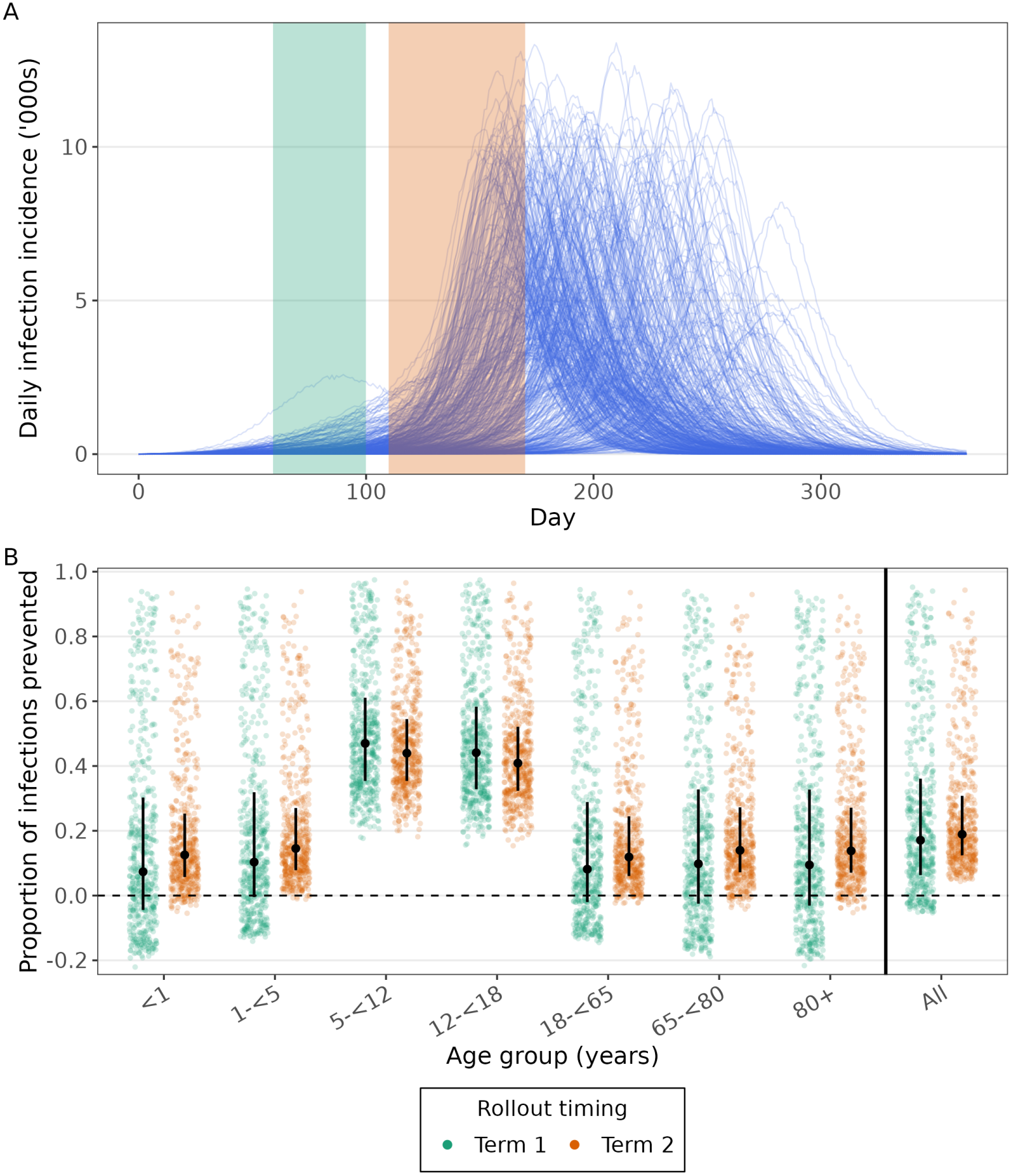
(**A**) Trajectories of daily infection incidence for the Baseline scenario (blue lines). The vaccination windows for term 1 (green, day 59–100) and term 2 (orange, day 110–170) are also shown. Term 1 vaccination occurred prior to the expected influenza season onset, while term 2 vaccination was during a period in which, for some model particles, infections were already increasing. (**B**) Comparison of the estimated impact of term 1 and term 2 LAIV rollout timings by age group. Proportion of infections prevented relative to Baseline under LAIV rollout in term 1 (green) and term 2 (orange), shown by age group and for the population overall. The shown results are for Central LAIV effectiveness, with 60% LAIV coverage in both primary and secondary school children (ages 5–*<*18). The horizontal dashed line indicates no change in infections relative to Baseline.

### 2.4 Impact of school-based LAIV programmes on clinical outcomes

The impact of a school-based LAIV programme on clinical outcomes depended on the assumed distribution of clinical outcomes by age. For each model particle, the total cumulative modelled infections at Baseline was used to compute age-stratified infection hospitalisation ratios (IHRs) according to a specified distribution of annual hospitalisations by age group. Those IHRs were then used in forward simulations to evaluate the impact of school-based LAIV programmes on hospitalisations.

We assumed an age distribution in hospitalisations based on Australian Institute of Health and Welfare data for 2023 (Figure S6). While the proportion of hospitalisations prevented by age group had a similar distribution to the proportion of infections prevented by age group (as expected), the proportion of hospitalisations prevented at the population-level was lower. This was because the target age groups (where we estimated the greatest impact on infections) have a lower rate of hospitalisation compared to non-target age groups. The absolute number of hospitalisations prevented by age group was dependent on the assumed distribution of annual hospitalisations by age. When a more severe influenza year/season (in terms of clinical burden) was used to calibrate the model, school-based LAIV programmes led to greater median reductions in overall hospitalisations. For example, under a 5–*<*18 rollout scenario of Central LAIV effectiveness and 40% coverage, we estimated median reductions in the absolute number of hospitalisations in ages 65–*<*80 per 100,000 people ranging from 25.8 during a ‘high’ burden year/season recorded by Australian jurisdictional influenza surveillance since 2020 (2024 in the state of Victoria) to 3.4 during a ‘low’ burden year/season (2023 in the state of New South Wales) (Figure 5). Additionally, the number of hospitalisations prevented per 100,000 people by LAIV effectiveness scenario (Figure S23) and the proportion of hospitalisations prevented per age group (Figure S14) are shown in the supplementary material.

**Figure 5:**
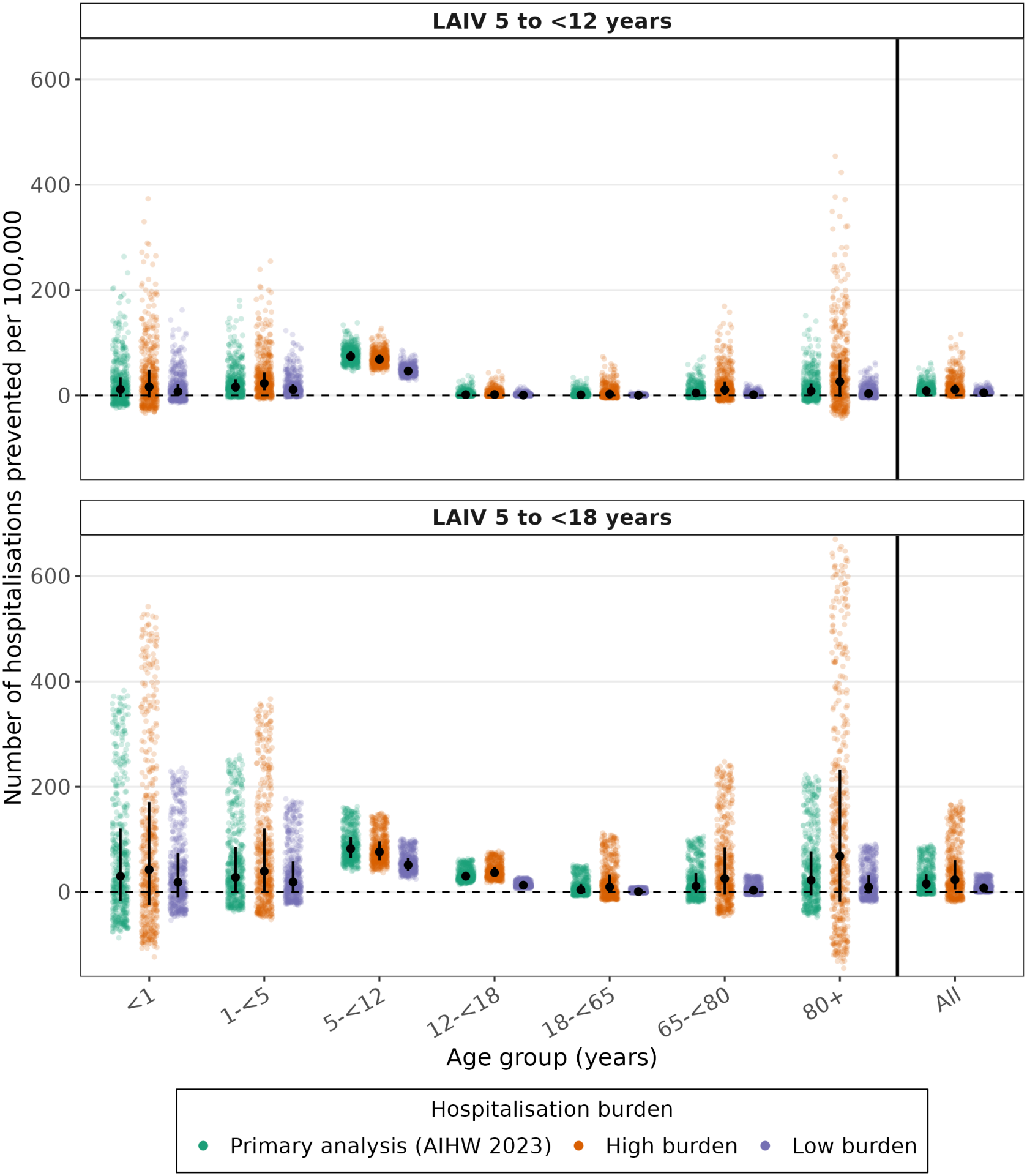
Estimated reduction in influenza-associated hospitalisations under school-based LAIV rollout (Central VE, 60% coverage), by age group and hospitalisation burden. Number of hospitalisations prevented per 100,000 population relative to the Baseline scenario for LAIV rollout targeting children aged 5–*<*12 years (top) and 5–*<*18 years (bottom). Estimates are shown using hospitalisation rates derived from AIHW 2023 data (green), a ‘high’ burden season (FluCAN data adjusted for 2024 in the state of Victoria) (orange), and a ‘low’ burden season (FluCAN data adjusted for 2023 in the state of New South Wales) (purple), by age group and for the population overall. The horizontal dashed line indicates no change in hospitalisations relative to the status quo. Negative values indicate an increase in hospitalisations.

## 3 Discussion

We have estimated the population-level and age-specific impact of school-based LAIV programmes in Australia under multiple scenarios for LAIV effectiveness and coverage, by developing a stochastic model of seasonal influenza transmission and calibrating it to data from Australian temperate regions. Overall, our findings suggest that school-based LAIV programmes could avert substantial influenza-associated health burden, including indirect effects in non-target age groups. While on average school-based LAIV programmes reduced infections and hospitalisations at the population level across all scenarios explored, effects varied across plausible transmission dynamics, particularly indirect effects. For some programme scenarios, we estimated that in non-target age groups both near complete suppression or a small increase in infections were possible outcomes. However, both of these outcomes were rare when LAIV was rolled out to school children in term 2 (rather than term 1 as in the primary analysis).

Limited previous modelling studies have estimated the impact of school-based LAIV programmes; the only existing model that we identified is described in Baguelin et al [16] (with the same model also used in Wenzel et al. [17] to perform a cost-effectiveness analysis). Our estimates exhibited a far greater level of uncertainty than Baguelin et al—likely due to differences in calibration processes (see below)—and were not directly comparable due to differences in the scenarios they considered. For example, compared to our analysis Baguelin et al considered different vaccination coverages in target age groups (15%, 30%, 50% and 70%), a slightly narrower target age group (5–16 year olds) and a different VE in working age adults (e.g. 70% in well matched years), and assumed that vaccines prevented infections only (i.e. no differences in severity for breakthrough infections). Additionally, Baguelin et al assumed that influenza vaccines provide ‘all-or-nothing’ protection, whereas we assume a vaccine that provides ‘leaky’ protection. While we calibrated vaccine effects in our model to recover the targeted vaccine effectiveness [18], the different mode of protection means that even if we had constructed the same vaccine coverage and effectiveness scenarios as Baguelin et al, we would not be able to directly compare outputs.

Most routine data collected to monitor trends in infection levels are inherently biased [19]. As such we calibrated our model to summary statistics using an Approximate Bayesian Computation approach, rather than calibrating the model to specific influenza time-series which would require an observation model with additional modelling assumptions. Baguelin et al [16] calibrated their model to age-stratified time series of subtype specific “ILI+” for England and Wales. While this surveillance indicator has been recommended for improved monitoring of trends in influenza infection incidence [20] it is not a true measure of infection incidence, and thus requires additional assumptions about the probability of symptoms and health-care/test seeking behaviour—that can vary by age and over time [21, 22]—to relate this data to modelled infection incidence. For Australia, we only had access to influenza case (daily, state-level) and hospitalisation (annual, national-level) data, which are subject to similar biases. We chose to calibrate our model to summary statistics on the expected infection dynamics in Australia to avoid making specific assumptions about the data generating processes, while incorporating a range of plausible infection dynamics that could explain the data. Accordingly, our results exhibited a high degree of uncertainty, reflecting general uncertainty in the underlying dynamics of influenza transmission. If data on age-stratified influenza infection incidence could be collected, more informative prior distributions and summary statistics could be defined and incorporated into our approach, improving the precision in our estimates, including the expected impacts on clinical outcomes.

The relative timing of underlying transmissibility (i.e. seasonality) and of realised epidemics was a key sensitivity on the estimated effects of a school-based LAIV programme. Influenza epidemics peak at similar times each year in temperate regions, with limited activity or growth during the summer months. Thus our model incorporated two mechanisms that act to modulate the timing of epidemic onset: seasonality in transmissibility and waning of pre-existing immunity. The model used in Baguelin et al. did not include either mechanism; as such, infections would begin increasing immediately at the start of the simulation and thus peak timing would be heavily controlled by the initial conditions and the basic reproduction number; depending on the initial conditions, this could lead to estimates of the reproduction number being biased downwards (upwards) to delay (bring forward) the epidemic peak when calibrating the model to data. In our main analysis, accepted model particles that resulted in increases in infections compared to the Baseline scenario had the following two properties: a peak in transmissibility (i.e. peak in seasonal forcing) late in the season; and a peak in infections (for the Baseline scenario) early in the season. However, it is unclear how common that combination of properties would be in reality. Theoretical models of recurrent infection dynamics have demonstrated that it is possible for peaks in transmissibility to lead or lag epidemics [23, 24], but their relative timings for influenza are likely sensitive to complex immune-evolutionary processes (and probably vary between years). Absolute humidity is considered to be a strong driver in the seasonality of influenza transmission [25], with lower levels of absolute humidity leading to higher levels of transmissibility; data of absolute humidity for Melbourne, Australia, suggests troughs in absolute humidity approximately consistent with the timings of peak transmissibility in our accepted model particles [26]. Our sensitivity analysis explored different assumptions about seasonal variation in transmissibility. When we assumed that the function varied less smoothly over time, i.e. more closely resembling a square wave than a sinusoidal function as in the primary analysis, we found fewer (but not zero) accepted parameter combinations led to increases in infections. While further exploration of the impact of modelling choices for seasonality is warranted, our current results indicate that the timing of school-based LAIV rollout is an important consideration for policymakers. We found that the timing of Baseline epidemics for model particles producing negative impacts in the main analysis (term 1 rollout) were not shifted as late in the season when LAIV was rolled out in term 2. This meant that projected epidemics under a term 2 rollout were less likely to coincide with the peak in transmissibility/seasonal forcing, resulting in fewer negative impacts and increasing the median estimated impact of the programme. Our potentially non-intuitive finding—where increased use of a transmission mitigating intervention can lead to overall higher disease burden—has been reported previously. Theoretical modelling studies investigating the effects of intervention timing in seasonally-forced transmission systems have reported model outcomes in which vaccination or treatment strategies (that reduce transmission) result in increased peak or final epidemic sizes [27, 28].

In this study we only considered a single influenza pathogen, when in reality the overall influenza burden each year can be composed of multiple influenza types/subtypes; though our estimates can still be interpreted as the effect on a single influenza subtype for a given year. In relating the effect on influenza infections to hospitalisations, we assumed that all allowed Baseline infection trajectories could explain the observed hospitalisation burden. In reality, variation in infection and hospitalisation burden between years would be correlated; in other words, when considering a year with more hospitalisations, we might expect parameter combinations that lead to greater numbers of infections to be more likely. We assumed that pre-existing immunity was uniform across age groups, but in reality we expect there to be some degree of heterogeneity. Patterns in heterogeneity may also vary between years. Age groups with lower relative levels of immunity will contribute relatively more to transmission, and so interventions targeting such age groups will likely be relatively more effective. It is unclear if school-age children will have higher or lower levels of pre-existing immunity: school-age children are younger and so will have accumulated fewer infections in their lifetime (i.e. less immunity), but higher contact rates will likely result in shorter times since the most recent infection (i.e. greater immunity).

Our model only considers influenza dynamics over a single year. Although our calibration procedure ensured that modelled infection dynamics were consistent with a typical influenza season, there can be substantial variation in influenza infection dynamics between years that our model was unable to capture. The effects of interventions were also only assessed over a single year. However, interventions in one year may influence dynamics in subsequent years due to resultant changes in the immune landscape. For example, lower attack rates due to interventions in one year could lead to greater levels of susceptibility (and attack rates) in following years [29]. While Bageulin et al. [16] considered multiple years of data, their model (like ours) only considered a single year in isolation. Predicting the ongoing impact of an annual school-based vaccination programme would require a model of the long-term dynamics of influenza, incorporating the effects of antigenic evolution and immunity (such as in [29]).

Overall, we found that high levels of uncertainty in underlying seasonal influenza transmission dynamics translated to a broad range of estimated outcomes under different school-based LAIV programme scenarios. Despite this uncertainty, we estimated that benefits of programme introduction could be substantial, including indirect effects in non-target age groups. We also showed that the timing of programme implementation may be important, with a later start to the programme (during the influenza season) found to mitigate against the risk of increased influenza infections in adults.

## 4 Methods

### 4.1 Methodological overview

We developed a stochastic, age-stratified SIR-type compartmental model of seasonal influenza transmission that incorporates vaccination and waning of pre-existing immunity (Figure 6). Within each age group, the population was separated into (mutually exclusive) compartments according to infection status, vaccination status, and the presence or absence of partial pre-existing immunity. Transitions between compartments represent the events of infection, recovery, vaccination, or waning of pre-existing immunity.

**Figure 6:**
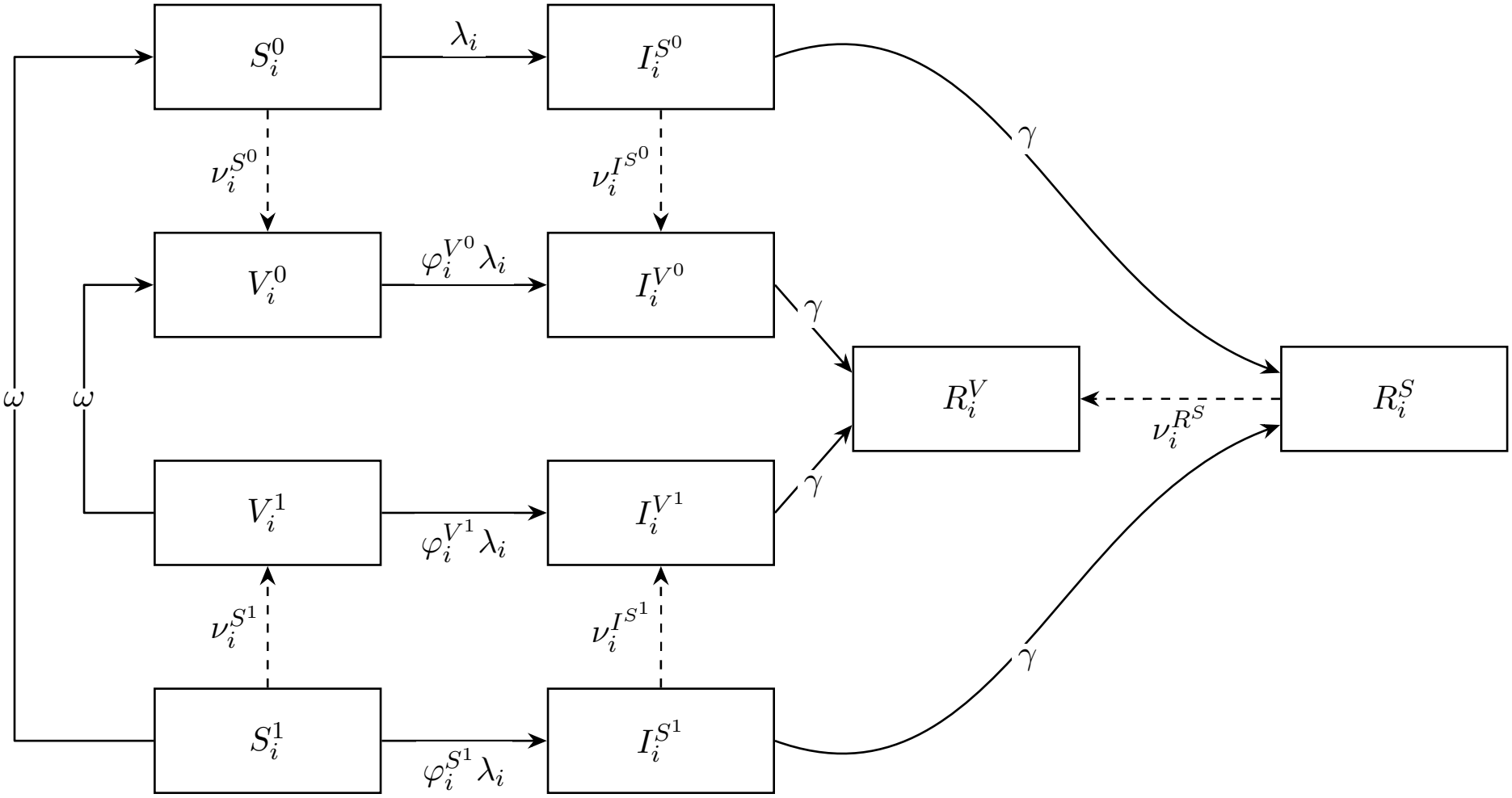
Schematic representation of the stochastic, compartmental, age-stratified influenza transmission model.

The model represents influenza transmission in a population with the age structure of Australia under a temperate seasonal transmission setting, producing a single annual winter influenza epidemic. Simulations were performed over a one-year period (January to December). Model parameter values consistent with the Australian context were identified using a combination of Approximate Bayesian Computation (ABC) and Latin Hypercube Sampling (LHS). The resulting posterior parameter sets were used to simulate a range of school-based live-attenuated influenza vaccine (LAIV) programmes under different assumptions regarding vaccine effectiveness (VE), vaccination coverage, and target age groups. For all scenarios, baseline inactivated influenza vaccine (IIV) programmes were assumed to be maintained in non-target age groups. The impact of introducing a school-based LAIV programme was evaluated by comparing epidemiological outcomes under each intervention scenario with a Baseline scenario representing current IIV vaccination coverage.

### 4.2 Model structure

The model stratifies the population by age group and epidemiological status. The ten epidemiological compartments considered include four susceptible compartments, four infectious compartments, and two recovered compartments. The susceptible compartments are 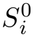 (fully susceptible), 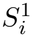 (susceptible with partial pre-existing immunity), 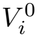 (vaccinated without pre-existing immunity), and 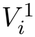 (vaccinated with partial pre-existing immunity). Each susceptible compartment has a corresponding infectious compartment, denoted 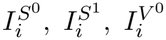, and 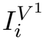. The two recovered compartments, 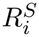 and 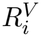, represent recovered, unvaccinated and vaccinated populations, respectively. The allowed transitions are shown as arrows in Figure 6.

Transitions between compartments (for a given age group, subscript *i*) are represented by the following system of ordinary differential equations, although the model was implemented stochastically as described in Section 4.3.

Susceptible compartment transitions:

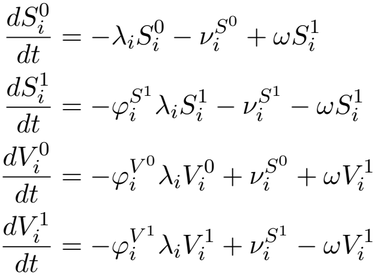

Infectious compartment transitions:

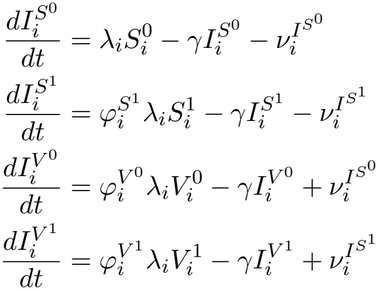

Recovered compartment transitions:

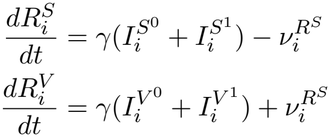

Omitting the age group subscript (*i*) and compartmental superscripts for clarity, *λ* is the force of infection (see below), *ν* is the rate of vaccine administration, *ω* is the waning rate of pre-existing immunity, *φ* is the relative susceptibility to infection, and *γ* is the recovery rate (see Table 1). Initial conditions for the model are described later under Model parametrisation and calibration (Section 4.4).

**Table 1:**
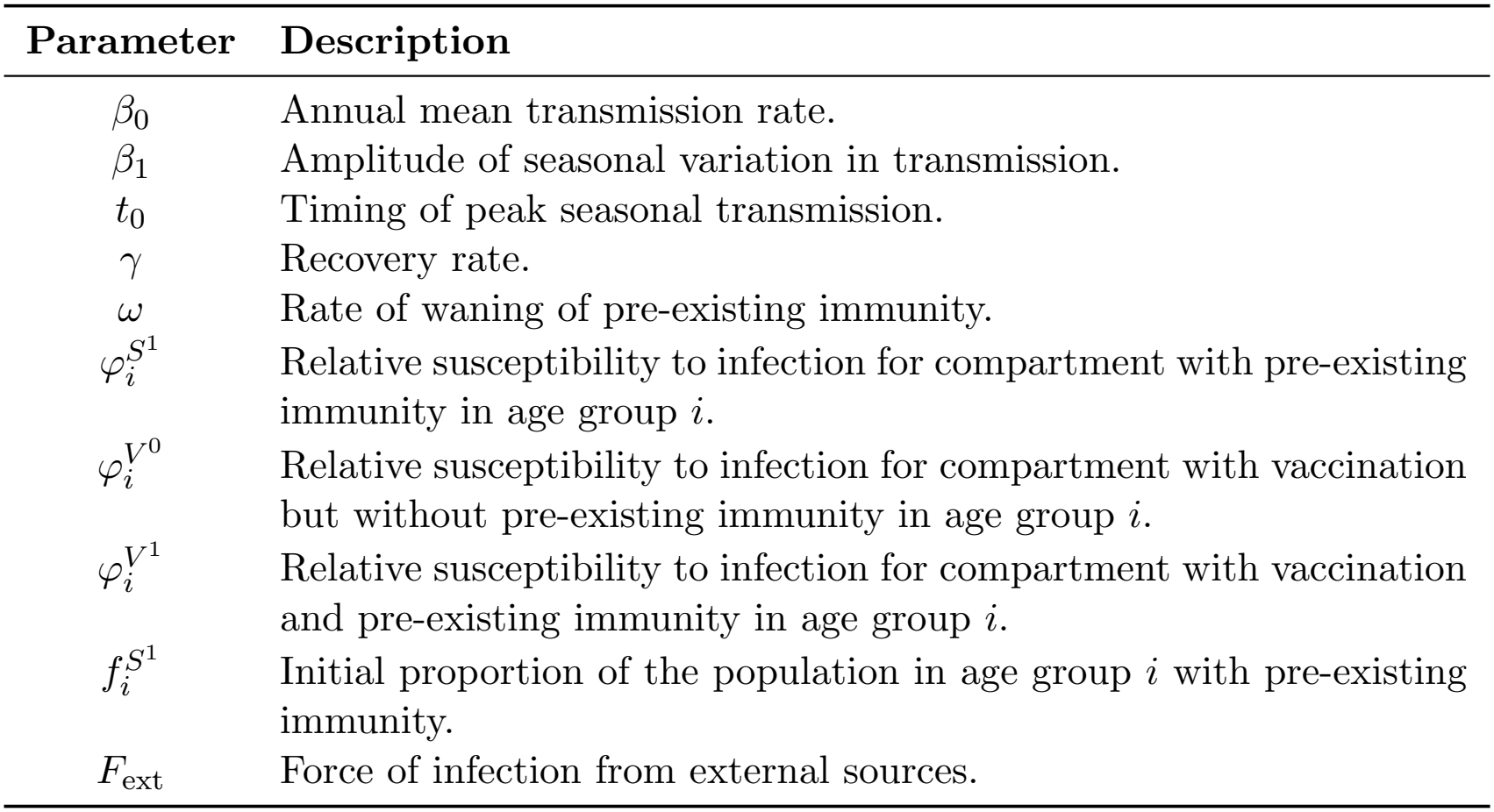
Model parameters.

| Parameter | Description |
|---|---|
| β<sub>0</sub> | Annual mean transmission rate. |
| β<sub>1</sub> | Amplitude of seasonal variation in transmission. |
| t<sub>0</sub> | Timing of peak seasonal transmission. |
| γ | Recovery rate. |
| ω | Rate of waning of pre-existing immunity. |
| φ<sub>i</sub><sup>S<sup>1</sup></sup> | Relative susceptibility to infection for compartment with pre-existing immunity in age group *i*. |
| φ<sub>i</sub><sup>V<sup>0</sup></sup> | Relative susceptibility to infection for compartment with vaccination but without pre-existing immunity in age group *i*. |
| φ<sub>i</sub><sup>V<sup>1</sup></sup> | Relative susceptibility to infection for compartment with vaccination and pre-existing immunity in age group *i*. |
| f<sub>i</sub><sup>S<sup>1</sup></sup> | Initial proportion of the population in age group *i* with pre-existing immunity. |
| F<sub>ext</sub> | Force of infection from external sources. |

We considered a population of 1, 000, 000, divided into eight age groups: *<* 1, 1–2, 3–4, 5–11, 12–17, 18–64, 65–79, and 80+ years. The proportion of the population in each age group was set to reflect the contemporary Australian population age distribution (Table 2). Because the simulation covered a single year, transitions between age groups (i.e. ageing) were not modelled, and thus the age distribution remained constant throughout the simulation. In the analyses of the outputs the age groups 1–2 and 3–4 were aggregated into one single group 1–4.

**Table 2:** Fraction of the population in each age group [31].

| **Age group** | **Fraction** |
|---|---|
| < 1 | 1.1% |
| 1–2 | 1.1% |
| 3–4 | 3.4% |
| 5–11 | 8.3% |
| 12–17 | 7.4% |
| 18–64 | 61.4% |
| 65–79 | 12.9% |
| 80+ | 4.4% |

The force of infection *λ*_*i*_ for age group *i* is given by:

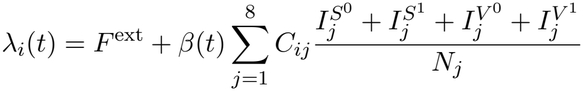

where

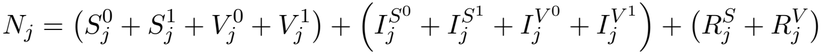

is the total population size of age group *j*. The external seeding term *F*^ext^ represents importation of infections from outside the modelled population. The time-varying transmission rate is given by

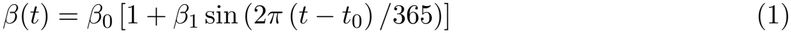

The time dependence in *β*(*t*) represents the seasonal variation in influenza transmission. The element *C*_*ij*_ of the contact matrix *C* denotes the average number of daily contacts that an individual in age group *i* has with age group *j*. We assumed that all infectious individuals contribute equally to transmission.

The effects of pre-existing immunity and vaccination are modelled as a reduction in susceptibility, relative to *S*^0^, controlled by the parameters 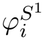 (unvaccinated with pre-existing immunity), 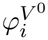 (vaccinated without pre-existing immunity), and 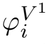 (vaccinated with pre-existing immunity). These range between 0 (complete immunity) to 1 (no change in susceptibility compared to *S*^0^). Vaccination is assumed to only reduce susceptibility to infection; our model does not allow for an effect on infectiousness, nor on the recovery rate following breakthrough infection.

The effect of LAIV was incorporated by modifying 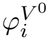 and 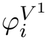 in the target age groups, while retaining their baseline values in all other age groups (effectively making the vaccination *φ* parameters age-dependent). That is, we did not model an immunisation programme in which an age group would have access to both LAIV and IIV vaccine products. Only target age groups for each of the LAIV intervention scenarios (see 4.5) were eligible to receive the LAIV. The IIV coverage and timing for all other age groups was unaffected by the LAIV intervention. Vaccines were administered daily throughout the rollout period, with protection assumed to be imparted immediately upon vaccination. Within each age group, the daily vaccination rate determined the overall transfer from unvaccinated to vaccinated compartments (susceptible, infectious and recovered), with doses allocated in proportion to the sizes of the respective unvaccinated compartments.

Our model allows for waning of pre-existing immunity in both unvaccinated (transitions from 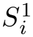 to 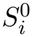) and vaccinated (transitions from 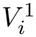 to 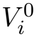) populations. Because our simulations covered a single influenza season, immunity acquired during the simulation (due to infection or vaccination) was assumed not to wane (i.e. there are no transitions from removed (*R*) compartments back to susceptible (*S*) or vaccinated compartments (*V*) to susceptible compartments).

### 4.3 Stochastic model dynamics

The underlying stochastic, continuous-time model was implemented using a tau-leaping algorithm [30], assuming daily time-steps (*τ* = 1 day). This algorithm approximates the continuous-time process by simulating multiple events over discrete time. For each transition rate *r*, the probability of a transition event occurring during a single time step was given by

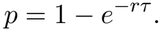

Infection probabilities were determined by the age-specific force of infection *λ*_*i*_, modified where appropriate by the susceptibility reduction factors 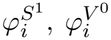, and 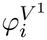. Recovery and waning probabilities were determined by the recovery rate *γ* and the waning rate *ω*, respectively. The recovery rate was the same for all ages and all four infectious compartments. Waning of pre-existing immunity occurred from *S*^1^ to *S*^0^ and from *V*^1^ to *V*^0^ at a constant rate, with the same rate across age groups.

At each time step, infection, recovery, and waning events were sampled using binomial random variables, and the resulting compartment changes were applied simultaneously.

Vaccines were administered at the start of each day, before infection, recovery, and waning events were simulated, provided the simulation was within the corresponding vaccination rollout period. For each age group, the daily number of vaccine doses was predetermined by the specified vaccination coverage and rollout schedule. These doses were allocated across all unvaccinated compartments in proportion to compartment size. Integer allocations were obtained using the largest remainder method: proportional allocations were first rounded down, and the remaining doses were assigned to the compartments with the largest fractional remainders, ensuring that the prescribed total number of doses was administered.

### 4.4 Model parametrisation and calibration

Model parameters were obtained from published literature where available, while the remaining parameters were estimated through calibration. Initial conditions for each simulation were specified as follows.

At the start of each simulation, all vaccinated, infectious, and recovered compartments were initialised to zero, such that the entire population was initially distributed between the 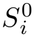 and 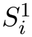 compartments. The fraction of the population initially assigned to 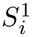 was determined by the parameter 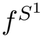.

The proportion of individuals in each age group was set to reflect the Australian population age distribution (Table 2). The fraction of the population with pre-existing immunity was assumed to be the same in all age groups.

The recovery rate was fixed at 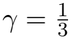, corresponding to a mean infectious period of 3 days. The external seeding parameter, *F*^ext^ was assumed to be constant over time and identical across age groups. Its magnitude was calibrated so that, in the absence of local transmission, external infections would result in approximately 0.1% of the population being infected over the one-year simulation period.

The contact matrix *C* (Figure S9) was computed using the POLYMOD survey data in the *R* package conmat [32], and national data on age group sizes from the Australian Bureau of Statistics [33].

Simulations were performed over a one-year period (1 January to 31 December). Model calibration was performed for the Baseline scenario, which included the existing IIV vaccination programme but no LAIV vaccination. The IIV campaign started on 1 March and was implemented over 24 weeks (see Figure S10). The rollout was parameterised using national weekly influenza vaccination coverage data by broad age group (6mo–*<*5, 5–*<*15, 15–*<*50, 50–*<*65, and *≥*65) provided by the National Centre for Immunisation Research and Surveillance (NCIRS) from 2020 to 31 May 2025 [34], with vaccines rolled out according to the mean vaccination rate from 2021–2025.

The model was calibrated to reproduce observed seasonal influenza dynamics in Australia. ABC was used to identify parameter combinations (model particles) that produced epidemic dynamics consistent with the calibration targets. The eight parameters considered were:*β*_0_, *β*_1_, *t*_0_, 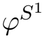, 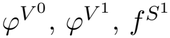, and *T*_1*/*2_. These parameters determine the annual mean transmission rate, the amplitude and timing of seasonal variation in transmission, relative susceptibility according to vaccination and pre-existing immunity status, the initial proportion of the population with pre-existing immunity, and the waning half-life of pre-existing immunity (with the waning rate *ω* = ln 2 */ T*_1*/*2_). We assumed uniform prior distributions, with their ranges summarised in Table 3.

**Table 3:**
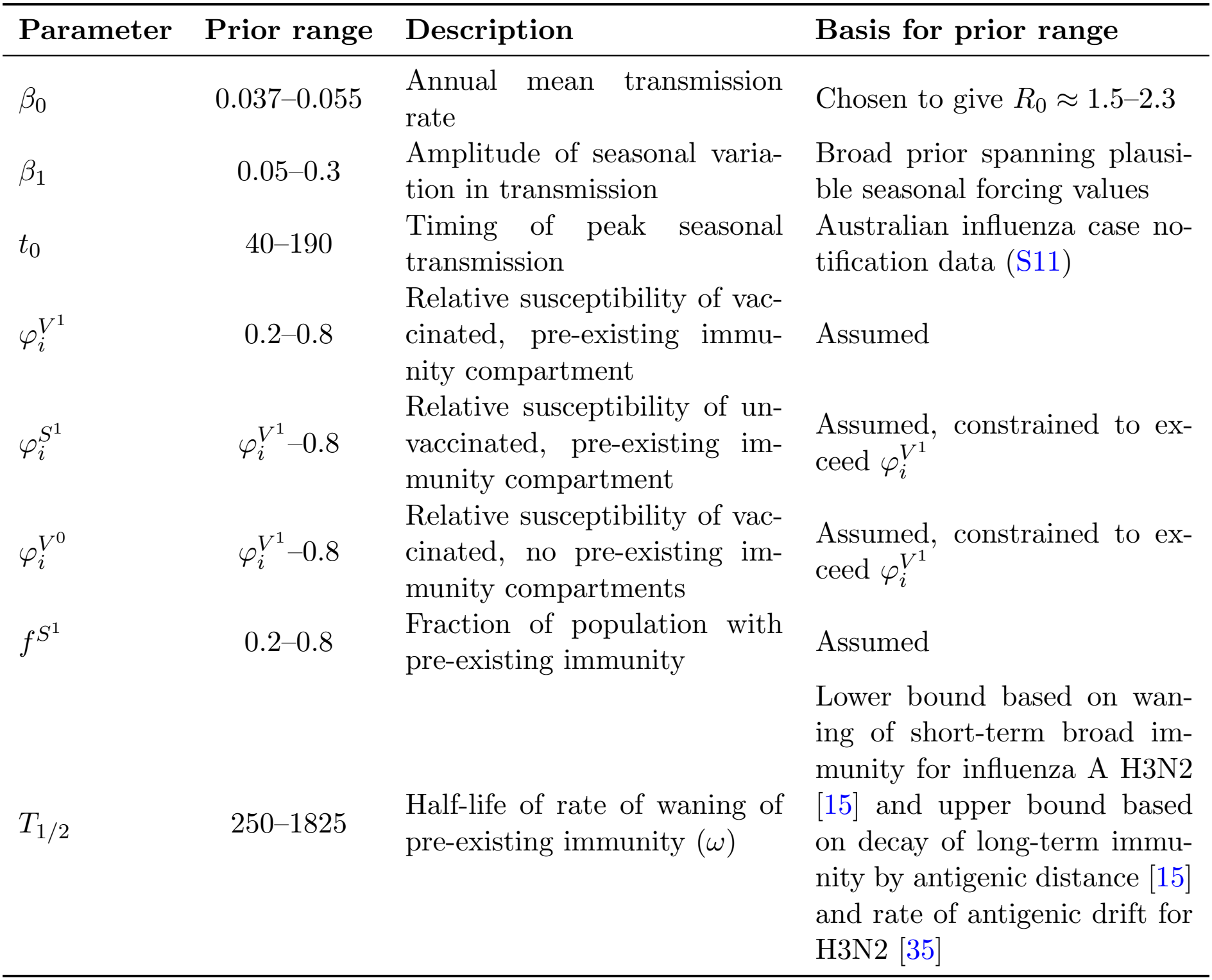
Prior ranges used for ABC calibration.

| Parameter | Prior range | Description | Basis for prior range |
|---|---|---|---|
| β<sub>0</sub> | 0.037–0.055 | Annual mean transmission rate | Chosen to give R<sub>0</sub> ≈ 1.5–2.3 |
| β<sub>1</sub> | 0.05–0.3 | Amplitude of seasonal varia-tion in transmission | Broad prior spanning plausi-ble seasonal forcing values |
| t<sub>0</sub> | 40–190 | Timing of peak seasonal transmission | Australian influenza case no-tification data (S11) |
| φ<sub>i</sub><sup>V<sup>1</sup></sup> | 0.2–0.8 | Relative susceptibility of vac-cinated, pre-existing immu-nity compartment | Assumed |
| φ<sub>i</sub><sup>S<sup>1</sup></sup> | φ<sub>i</sub><sup>V<sup>1</sup></sup>–0.8 | Relative susceptibility of un-vaccinated, pre-existing im-munity compartment | Assumed, constrained to ex-ceed φ<sub>i</sub><sup>V<sup>1</sup></sup> |
| φ<sub>i</sub><sup>V<sup>0</sup></sup> | φ<sub>i</sub><sup>V<sup>1</sup></sup>–0.8 | Relative susceptibility of vac-cinated, no pre-existing im-munity compartments | Assumed, constrained to ex-ceed φ<sub>i</sub><sup>V<sup>1</sup></sup> |
| f<sup>S<sup>1</sup></sup> | 0.2–0.8 | Fraction of population with pre-existing immunity | Assumed |
| T<sub>1/2</sub> | 250–1825 | Half-life of rate of waning of pre-existing immunity (ω) | Lower bound based on wan-ing of short-term broad im-munity for influenza A H3N2 [15] and upper bound based on decay of long-term immu-nity by antigenic distance [15] and rate of antigenic drift for H3N2 [35] |

The prior range for *β*_0_ was chosen to give a basic reproduction number of approximately *R*_0_ = 1.5–2.3 [36], where *R*_0_ was calculated using the annual mean transmission rate *β*_0_. The remaining prior ranges were selected to span biologically plausible values while remaining intentionally broad. Parameter values were sampled from uniform prior distributions using LHS, which stratifies each prior distribution and ensures efficient coverage of the parameter space. For 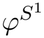 and 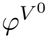, we imposed the constraints 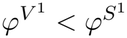 and 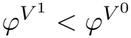, reflecting the assumption that hybrid immunity provides greater protection than either vaccination or prior infection alone.

Model calibration was performed using a simple rejection-sampling ABC algorithm. Recently, Vollert et al. [37] formalised the interpretation of ABC as assessing consistency between a model and a set of beliefs informed by empirical data, expert knowledge, and theoretical considerations. Following this framework, we defined a set of calibration targets representing characteristic features of seasonal influenza epidemics in Australia.

We sampled 70, 000 candidate parameter vectors from the prior distributions using LHS. For each parameter vector, ten stochastic realisations were simulated. Three summary statistics were calculated for each realisation. A parameter vector was accepted if at least nine of the ten realisations fell within the acceptance ranges for all three summary statistics, otherwise it was rejected. Requiring agreement across multiple stochastic realisations reduced the probability of accepting parameter vectors that matched the calibration targets purely because of stochastic variability.

Calibration was based on three summary statistics: the population-level infection attack rate, epidemic peak timing, and seasonal VE. Their target ranges are summarised in Table 4.

**Table 4:** ABC targets, the range of target values, and their source.

| **ABC target** | **Range** | **Source** |
|---|---|---|
| Population-level infection attack rate | 10–45% | [15, 38] |
| Seasonal vaccine effectiveness against infection | 38—50% | Annual Australian Respiratory Surveillance Report [9] |
| Peak timing | 147–289 | Analysis of Australian influenza case data (see Section 4.4 and Figure S11). |

The target range for the population-level infection attack rate is based on [15, 38].

To establish a range of target values for peak timing, we analysed state-level time series of daily influenza cases from the Nationally Notifiable Diseases Surveillance System (NNDSS). Data from the 2015–2019 and 2022–2025 influenza seasons were included, excluding the period during which COVID-19 restrictions substantially disrupted influenza transmission. All Australian states and territories were included except the Northern Territory, whose influenza dynamics are not characterised by strong seasonal forcing. For each season and jurisdiction, we identified the day of maximum daily influenza case incidence. The mean peak day was 218 with a standard deviation of 36 days (see Figure S11). The ABC acceptance range for peak timing was defined as the mean *±*1.96 standard deviations.

The target range for the IIV VE was based on the Annual Australian Respiratory Surveillance Report [9]. For each realisation, the VE was calculated using the attack rate of the vaccinated population *AR*_vacc_ and that of the unvaccinated population *AR*_unvacc_.

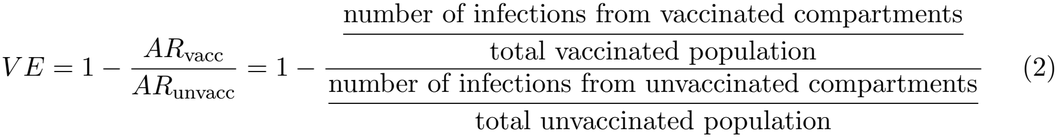

This definition represents the relative reduction in infection attack rate among the vaccinated compared with the unvaccinated. This approach for calibrating the effect of vaccination in epidemic models to estimates of VE has been recommended by Middleton et al. [18].

Because the model allows vaccination after infection, the standard VE calculation required adjustment. Only infections occurring after vaccination contributed to the vaccinated attack rate, whereas infections occurring before vaccination contributed to the unvaccinated attack rate. Vaccinations administered after infection were therefore tracked throughout each simulation and excluded from the vaccinated denominator, with an analogous correction applied to the unvaccinated denominator.

### 4.5 Simulated intervention scenarios

Following model calibration, accepted parameter vectors were used to simulate multiple scenarios for a school-based LAIV programme. The simulated scenarios differed along three possible axes: the target age group for the intervention (either primary school only (5 *™ ™ <* 11) or including secondary school as well (5 *™ ™ <* 18)); the coverage of the LAIV within the target age group (20%, 40%, 60%, or 80%); and the VE against infection (categorised as Pessimistic (VE *∈* [0.38, 0.50]), Central (VE *∈* [0.44, 0.69]), or Optimistic (VE *∈* [0.63, 0.88]) within the target age group. The LAIV campaigns were implemented over a six week period starting on March 1 (day 59–100, consistent with rollout during term 1 of the school year) with vaccines administered at a constant rate (see Figure S10). We also considered a sensitivity analysis in which the LAIV campaign was implemented over a 9 week period starting on April 20 (day 110–170, consistent with a rollout during term 2 of the school year). The effects of each intervention scenario were compared with the Baseline scenario, in which no LAIV intervention occurred.

For each accepted particle, new values for 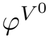 and 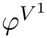 were generated representing susceptibility reductions due to LAIV (as opposed to IIV, that were obtained in the calibration step). Age groups that did not receive LAIV retained the original *φ* values from the calibration step, effectively making 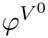 and 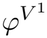 age-dependent. The new *φ* values (for LAIV) were generated by an additional ABC step. For this step, simulations used Baseline vaccination coverage, with LAIV replacing IIV for ages 5–*<* 18 years. VE was calculated using Equation 2, restricted to the 5–*<* 18 year age groups receiving LAIV. The target range for the VE in this additional ABC step were the same as the ones mentioned previously: Pessimistic (VE *∈* [0.38, 0.50]), Central (VE *∈* [0.44, 0.69]), or Optimistic (VE *∈* [0.63, 0.88]). The VE was calculated using definition (2) restricted to the age groups receiving LAIV. In this additional ABC step, we also imposed the constraints 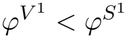 and 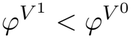, such that the new LAIV-associated values retained the assumption that hybrid immunity provides greater protection than either prior infection or vaccination alone.

For each accepted particle we simulated 50 influenza epidemics for the Baseline scenario and for each intervention scenario. For a given particle, the outcomes of each simulation (e.g. total infections by age group) for an intervention scenario were compared with the outcomes of simulations of the Baseline scenario (randomly paired), yielding 50 estimates of the intervention’s effect. The mean of these 50 estimates was then calculated for each parameter set, resulting in one estimate of the intervention effect per accepted particle. This pairing ensured that intervention and Baseline outcomes were compared using the same underlying parameter combinations.

### 4.6 Mapping infections to hospitalisations

Hospitalisations were not modelled explicitly within the transmission model, but were calculated from simulated infections using age-specific infection hospitalisation rates (IHRs). For each age group, we specified a target annual number of influenza hospitalisations, *H*_*i*_. For each accepted particle (i.e. a set of parameters), an age-specific IHR was calculated such that the Baseline model output reproduced these target hospitalisation numbers.

For each accepted particle, the mean numbers of infections across the 50 Baseline realisations were calculated by age group and vaccination status, denoted 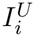 for the unvaccinated and 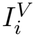 for the vaccinated. The age specific IHR was then calculated as

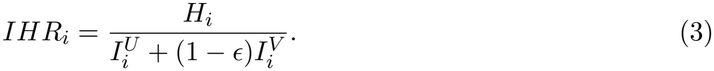

VE in the transmission model was defined as protection against infection. Protection against hospitalisation conditional on infection was incorporated separately through *ϵ*, such that vaccinated individuals who became infected had a relative probability 1 *™ ϵ* of hospitalisation compared with infected unvaccinated individuals. We assumed *ϵ* = 0.2 [39].

The target age-specific annual hospitalisation numbers *H*_*i*_ were based on national influenza hospitalisation rates per 100,000 population for 2023. These were calculated using the July–December and January–June data from the 2022–23 and 2023–24 AIHW Principal Diagnosis Data Cubes, respectively, filtered to codes J09, J10, J10.1, and J10.8, and linked to ABS population data for 2023.

Age-specific IHRs were assumed to remain unchanged across intervention scenarios. Hospitalisations for each intervention scenario were therefore calculated from the corresponding simulated numbers of vaccinated and unvaccinated infections as

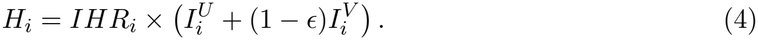

As a sensitivity analysis we also considered how our results (i.e. absolute number of hospitalisations prevented by age group) would be affected if the annual hospitalisation rates per 100,000 were typical of a ‘high’ or ‘low’ hospitalisation burden year. The AIHW data used in our main analysis was unavailable for other years; instead we used hospitalisation data by year and jurisdiction from FluCAN [REF] to select rates to represent a ‘high’ and ‘low’ hospitalisation burden year. Data was only included from 2023 and 2024 (earlier data was influenced by the COVID-19 pandemic, and 2025 was not available at the time of analysis). FluCAN hospitalisation numbers are likely not representative of the overall hospitalisation burden due to low coverage (approximately 1/6 and 1/2 of hospital beds for adults and children respectively). As such we calculated the multiplicative difference by age group between rates in the AIHW 2023 data and in the FluCAN data nationally for 2023. We then scaled the FluCAN rates for jurisdiction and year combinations representing a ‘high’ and ‘low’ burden year using these multiplicative differences. The ‘high’ and ‘low’ burden jurisdiction/year were selected based on their overall hospitalisation rates, considering only states with populations greater than one million people (as smaller jurisdictions were likely to have greater variance in FluCAN’s coverage). The ‘high’ burden year was for Victoria in 2024, and the ‘low’ burden year was for New South Wales in 2023 (Figure S6). These two years likely do not accurately represent the true variation in hospitalisation rates in Australia between years (due to the small sample size), but offer a representative example as to how our results will vary based on the overall hospitalisation burden used for calibration.

## Data Availability

Aggregate hospitalisation data from the Australian Institute of Health and Welfare (AIHW), population data by jurisdiction from the Australian Bureau of Statistics (ABS), and a CSV file containing influenza epidemic peak timings are publicly available in the GitHub repository, as well as the code used to implement the model, run the simulations, and generate the analyses presented in this study.
The peak timing were derived from influenza case data from the Australian National Notifiable Disease Surveillance System (NNDSS), which are not able to be shared under the terms of our data access agreements. For access to NNDSS data, a request must be submitted via which will be assessed by a data committee independent of the authorship group.

https://github.com/DrBoxie/ACEFA_scenario_modelling

## Acknowledgements

This research is supported by the Australia–Aotearoa Consortium for Epidemic Forecasting and Analytics (ACEFA), a National Health and Medical Research Council of Australia Centre of Research Excellence (2035303). OE is supported by a University of Melbourne McKenzie fellowship. FMS is supported by the National Health and Medical Research Council of Australia through the Investigator Grant Scheme (Emerging Leader Fellowship, 2021/GNT2010051). JMM is supported by the Australian Research Council through the Laureate Fellowship Scheme (FL240100126).

National Notifiable Disease Surveillance System data were provided by the Interim Australian Centre for Disease Control, Australian Government Department of Health and Aged Care, on behalf of the Communicable Diseases Network Australia (CDNA). We thank public health staff and data managers in state and territory health departments, and the Australian Government Department of Health and Aged Care, along with state and territory public health laboratories.

This research was supported by use of the Nectar Research Cloud, a collaborative Australian research platform supported by the NCRIS-funded Australian Research Data Commons (ARDC), and by the University of Melbourne Research Computing Services.

The authors wish to acknowledge efforts of the ACEFA Hub Infrastructure Team in supporting the Australia–Aotearoa Scenario Modelling Hub initiative (Katharine Senior, Ruarai Tobin, and Alys Young) and helpful comments from the University of New South Wales modelling team (Tiffany Leung, Isaac Stopard and James Wood) and ACEFA reviewers (Michael Plank, Nick Golding, and Kathtryn Glass) during transmission model development.

## Data and code availability

Aggregate hospitalisation data from the Australian Institute of Health and Welfare (AIHW), population data by jurisdiction from the Australian Bureau of Statistics (ABS), and a CSV file containing influenza epidemic peak timings are publicly available in the GitHub repository, as well as the code used to implement the model, run the simulations, and generate the analyses presented in this study.

The peak timing were derived from influenza case data from the Australian National Notifiable Disease Surveillance System (NNDSS), which are not able to be shared under the terms of our data access agreements. For access to NNDSS data, a request must be submitted via which will be assessed by a data committee independent of the authorship group.

