## Supplemental material for "Modelling the impact of a school-based influenza vaccination programme under uncertainty in seasonal transmission dynamics"

### Supplementary Materials

| Scenario VE | LAIV coverage | LAIV target | Median % infection reduction | IQR % infection reduction | 95% CrI % infection reduction |
|---|---|---|---|---|---|
| Pessimistic | 20% | 5–<12 | 0.96 | 0.15 – 2.24 | -0.70 – 8.55 |
| Pessimistic | 40% | 5–<12 | 2.76 | 1.11 – 5.58 | -0.78 – 19.68 |
| Pessimistic | 60% | 5–<12 | 4.66 | 2.35 – 9.06 | 0.71 – 30.69 |
| Pessimistic | 80% | 5–<12 | 6.98 | 3.75 – 12.47 | -0.29 – 41.71 |
| | | | | – | |
| Pessimistic | 20% | 5–<18 | 1.25 | -0.57 – 4.60 | -2.40 – 22.27 |
| Pessimistic | 40% | 5–<18 | 4.91 | 0.33 – 13.14 | -4.28 – 57.05 |
| Pessimistic | 60% | 5–<18 | 10.21 | 2.46 – 25.06 | -5.45 – 81.48 |
| Pessimistic | 80% | 5–<18 | 17.72 | 6.46 – 40.37 | -5.79 – 91.02 |
| | | | | – | |
| Central | 20% | 5–<12 | 1.69 | 0.57 – 3.24 | -0.47 – 11.33 |
| Central | 40% | 5–<12 | 4.15 | 1.88 – 7.30 | -0.23 – 24.23 |
| Central | 60% | 5–<12 | 6.80 | 3.67 – 11.63 | 0.30 – 36.19 |
| Central | 80% | 5–<12 | 9.79 | 5.53 – 15.95 | 1.19 – 47.45 |
| | | | | – | |
| Central | 20% | 5–<18 | 2.63 | -0.09 – 6.67 | -2.48 – 29.61 |
| Central | 40% | 5–<18 | 8.09 | 2.15 – 18.74 | -3.60 – 68.06 |
| Central | 60% | 5–<18 | 17.10 | 6.34 – 36.08 | -3.77 – 88.63 |
| Central | 80% | 5–<18 | 29.35 | 12.49 – 55.29 | -2.48 – 94.56 |
| | | | | – | |
| Optimistic | 20% | 5–<12 | 2.68 | 1.29 – 4.77 | -0.17 – 15.02 |
| Optimistic | 40% | 5–<12 | 6.35 | 3.66 – 10.74 | 0.67 – 32.38 |
| Optimistic | 60% | 5–<12 | 9.97 | 6.31 – 16.44 | 1.93 – 44.91 |
| Optimistic | 80% | 5–<12 | 13.61 | 8.98 – 21.54 | 3.26 – 57.19 |
| | | | | – | |
| Optimistic | 20% | 5–<18 | 4.67 | 1.60 – 10.80 | -2.07 – 39.41 |
| Optimistic | 40% | 5–<18 | 14.04 | 5.98 – 29.36 | -2.48 – 81.49 |
| Optimistic | 60% | 5–<18 | 26.21 | 13.12 – 56.11 | -1.35 – 94.02 |
| Optimistic | 80% | 5–<18 | 40.64 | 22.17 – 76.29 | 1.59 – 96.59 |

Table S1: Reduction in infections compared to Baseline across all studied scenarios (for $K = 0$, i.e. the sine wave seasonal forcing).

| **Scenario VE** | **LAIV coverage** | **LAIV target** | **Median % infection reduction** | **IQR % infection reduction** | **95% CrI % infection reduction** |
|---|---|---|---|---|---|
| Pessimistic | 20% | 5–<12 | 1.53 | 0.65 – 2.85 | -0.29 – 9.18 |
| Pessimistic | 40% | 5–<12 | 4.18 | 2.45 – 7.19 | 0.41 – 21.33 |
| Pessimistic | 60% | 5–<12 | 7.00 | 4.53 – 11.52 | 1.15 – 33.39 |
| Pessimistic | 80% | 5–<12 | 10.05 | 6.80 – 15.84 | 2.32 – 44.59 |
| | | | | – | |
| Pessimistic | 20% | 5–<18 | 1.28 | -0.61 – 4.78 | -2.53 – 22.53 |
| Pessimistic | 40% | 5–<18 | 5.25 | 0.50 – 13.71 | -4.29 – 57.24 |
| Pessimistic | 60% | 5–<18 | 11.03 | 2.75 – 26.07 | -5.11 – 81.57 |
| Pessimistic | 80% | 5–<18 | 18.75 | 6.99 – 41.58 | -5.37 – 90.91 |
| | | | | – | |
| Central | 20% | 5–<12 | 2.30 | 1.15 – 3.94 | 0.022 – 12.32 |
| Central | 40% | 5–<12 | 5.84 | 3.42 – 9.28 | 1.13 – 26.12 |
| Central | 60% | 5–<12 | 9.43 | 5.96 – 14.51 | 2.41 – 38.61 |
| Central | 80% | 5–<12 | 13.31 | 8.62 – 19.58 | 3.92 – 50.25 |
| | | | | – | |
| Central | 20% | 5–<18 | 2.50 | -0.32 – 6.81 | -2.85 – 29.72 |
| Central | 40% | 5–<18 | 8.09 | 2.07 – 19.24 | -4.018 – 68.55 |
| Central | 60% | 5–<18 | 17.08 | 6.51 – 37.00 | -4.26 – 88.41 |
| Central | 80% | 5–<18 | 29.35 | 12.88 – 55.74 | -2.97 – 94.34 |
| | | | | – | |
| Optimistic | 20% | 5–<12 | 3.50 | 2.05 – 5.79 | 0.51 – 16.16 |
| Optimistic | 40% | 5–<12 | 8.32 | 5.45 – 12.82 | 2.23 – 34.30 |
| Optimistic | 60% | 5–<12 | 12.99 | 9.11 – 19.59 | 4.35 – 47.63 |
| Optimistic | 80% | 5–<12 | 17.42 | 12.61 – 25.61 | 6.66 – 60.06 |
| | | | | – | |
| Optimistic | 20% | 5–<18 | 4.39 | 1.07 – 10.76 | -2.78 – 39.43 |
| Optimistic | 40% | 5–<18 | 13.90 | 5.27 – 29.67 | -3.30 – 81.47 |
| Optimistic | 60% | 5–<18 | 26.45 | 12.95 – 56.57 | -2.24 – 93.75 |
| Optimistic | 80% | 5–<18 | 41.10 | 21.50 – 76.57 | 1.06 – 96.32 |

Table S2: Reduction in hospitalisations compared to Baseline across all studied scenarios (for $K = 0$, i.e. the sine wave seasonal forcing).

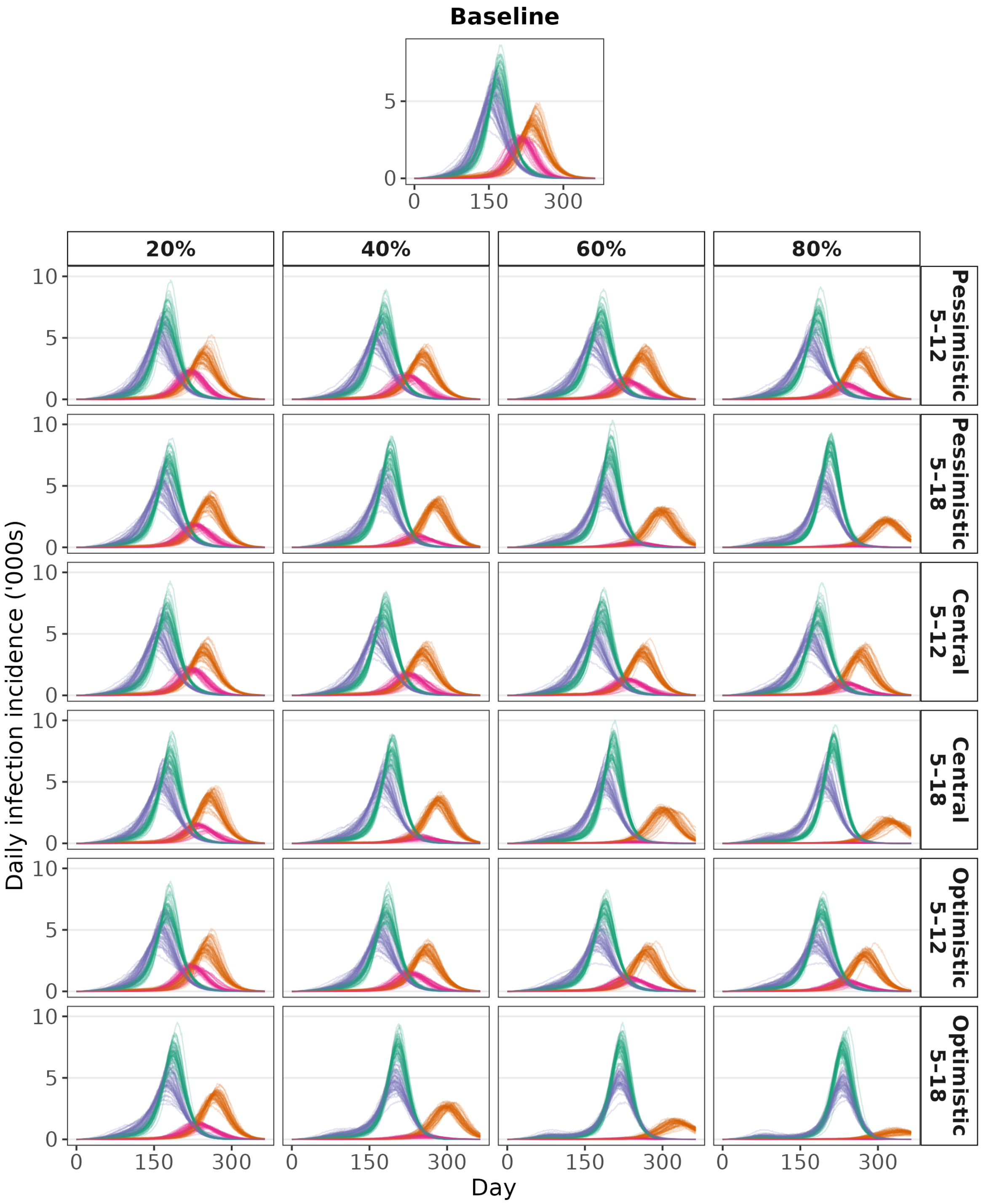


Figure S1: Daily infection incidences for 4 randomly selected particles across all scenarios. For each particle, indicated by colour, all 50 stochastic simulations are shown. The clustering of trajectories by colour illustrates that variation in epidemic dynamics is driven primarily by differences between particles rather than stochastic variation between runs (i.e. differences in the random seed). For the intervention scenarios, columns indicate LAIV coverage in the target age group (20%, 40%,60%, or 80%), while rows indicate the combination of target age groups (5–<12 or 5–< 18) and VE assumption (Pessimistic, Central, or Optimistic). The Baseline scenario is shown separately above the intervention scenarios.

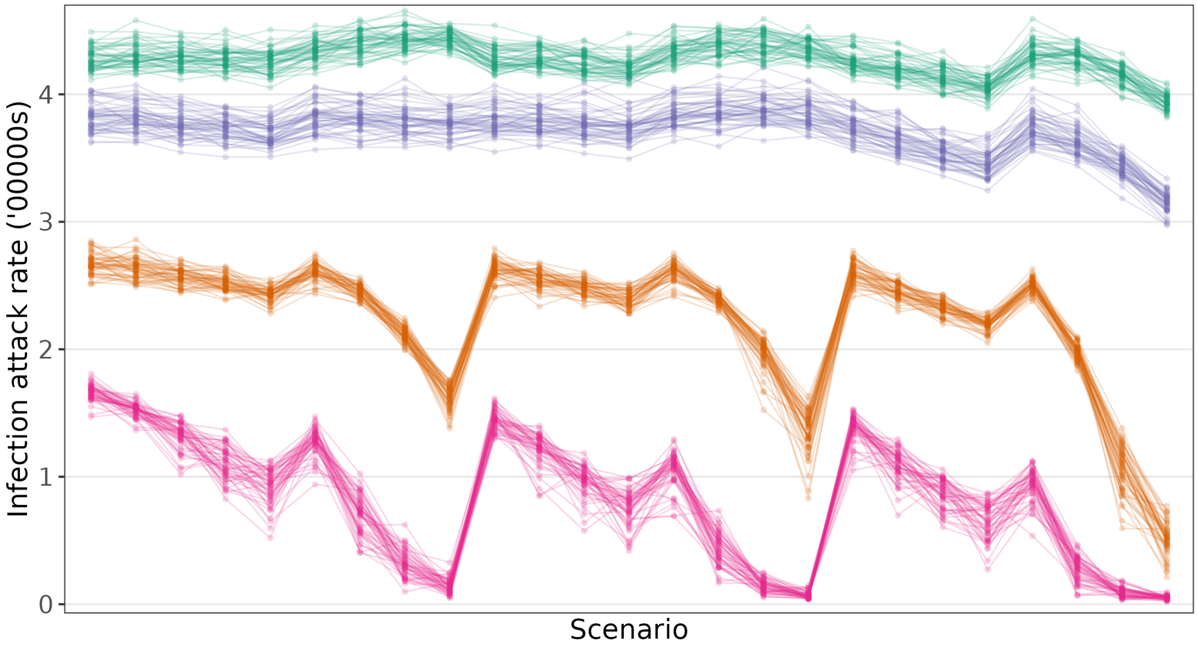


Figure S2: Infection attack rates across all scenarios for 4 randomly selected particles. For each selected particle (i.e. each colour on the plot), the results of all 50 runs are shown. This illustrates that differences in attack rates are mostly driven by differences in the particles themselves rather than by stochastic effects (i.e. the initial random seed).

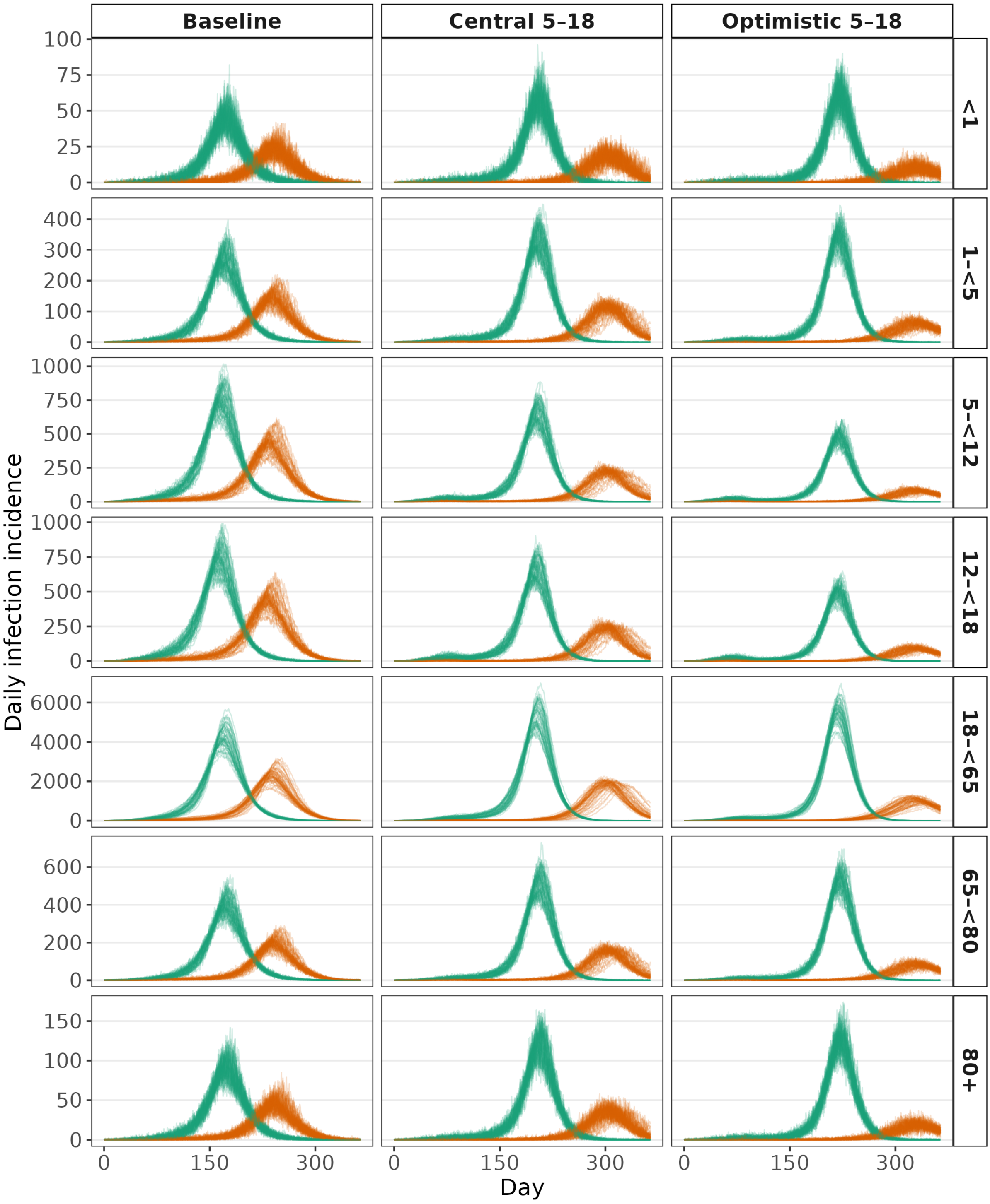


Figure S3: Daily infection incidences for 2 randomly selected particles across 3 of the scenarios, stratified by age group. For each selected particle (i.e. each colour on the plot), the results of all 50 runs are shown. This illustrates that differences in daily infection incidence are mostly driven by differences in the particles themselves rather than by stochastic effects (i.e. the initial random seed).

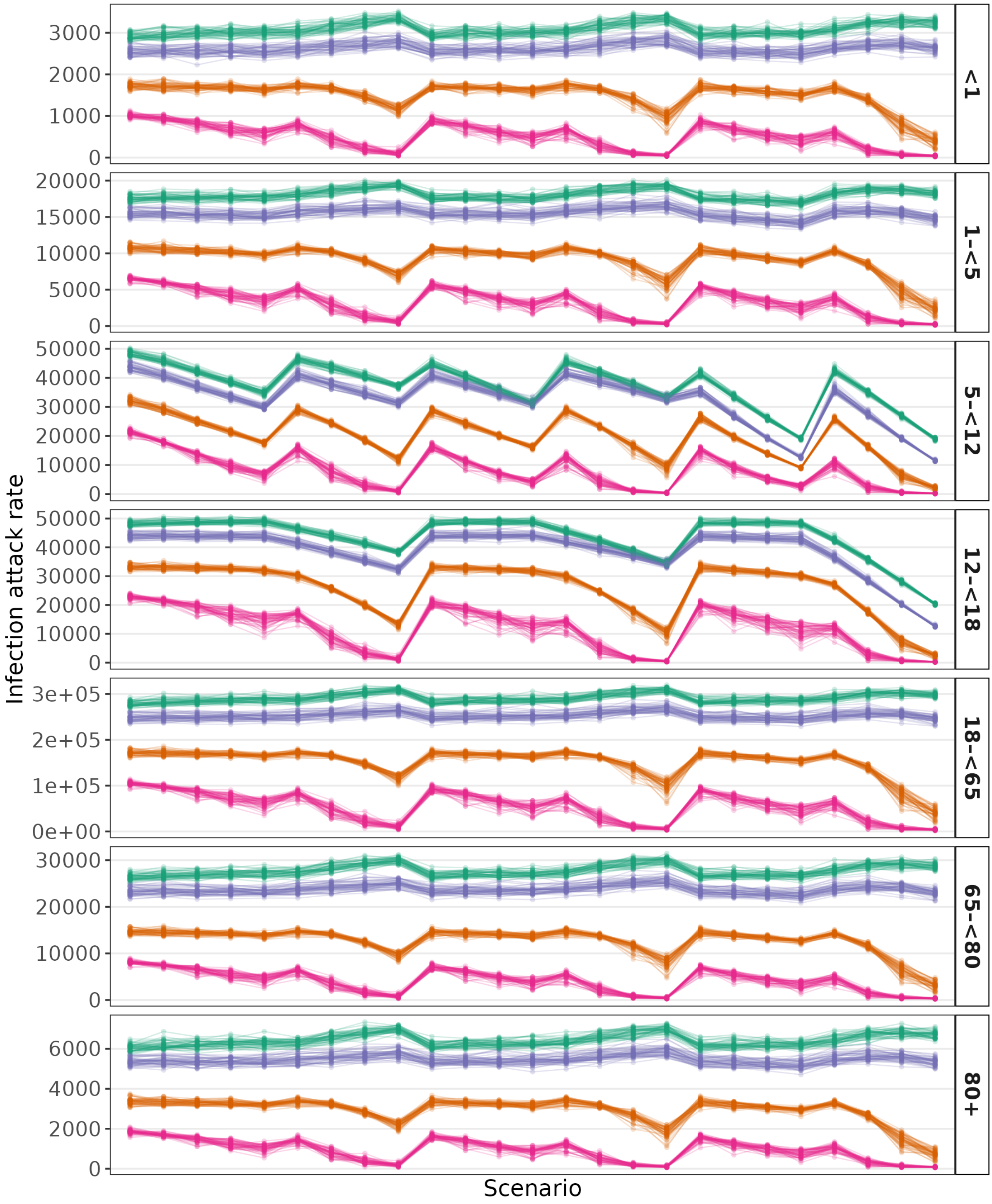


Figure S4: Infection attack rates across all scenarios for 4 randomly selected particles stratified by age group. Each position on the horizontal axis represents a different simulation scenario. Their labels are omitted for visual clarity. For each selected particle (i.e. each colour on the plot), the results of all 50 runs are shown. This illustrates that differences in attack rates for each age group are mostly driven by differences in the particles themselves rather than by stochastic effects (i.e. the initial random seed).

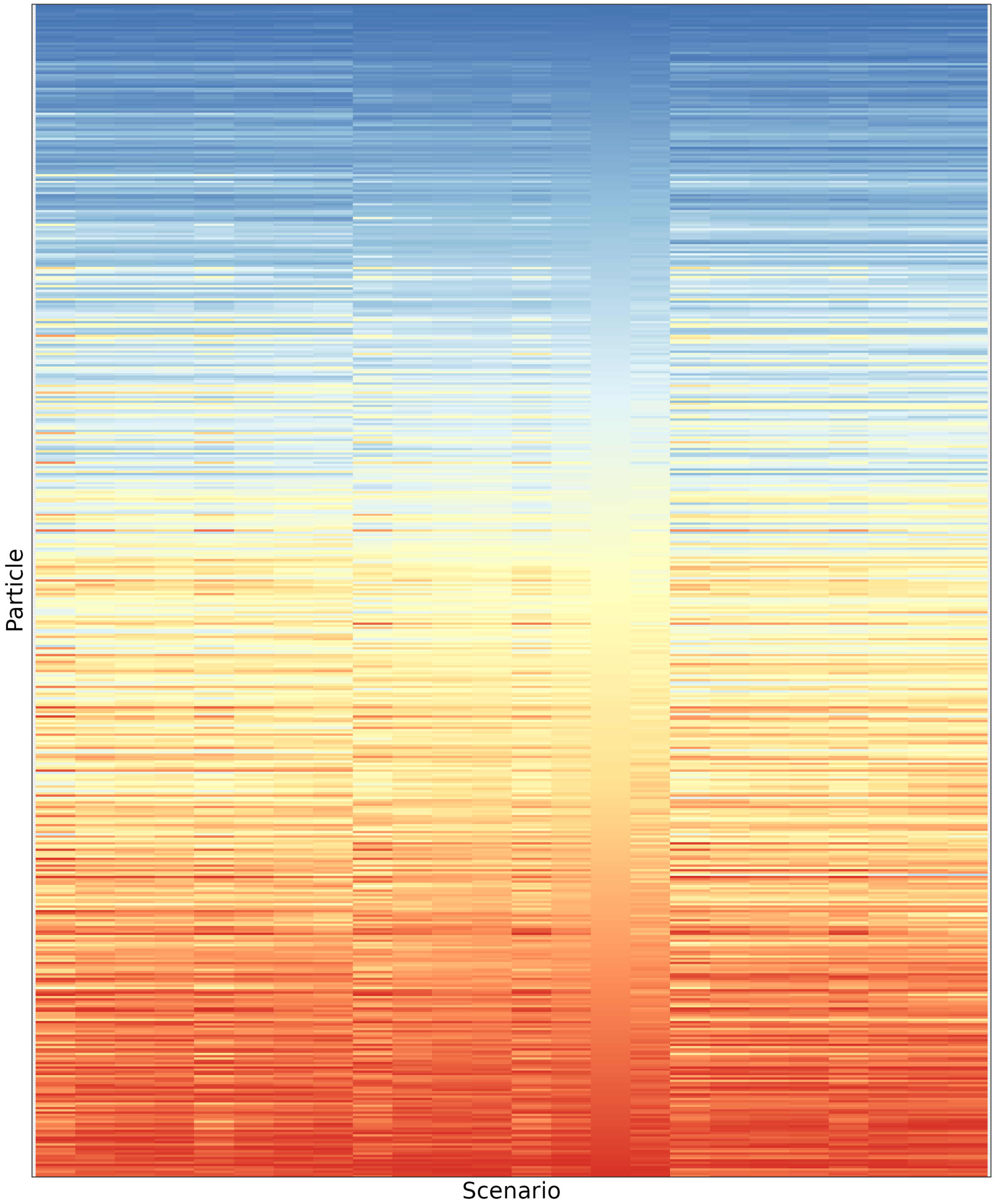


Figure S5: Particle rankings by mean infection reduction with respect to Baseline. Each row of the heat map represents one of the 518 particles accepted in the ABC step. Each column represents one of the 24 (non-Baseline) scenarios. The colour represents the rank when sorted from most reduction in infections with respect to Baseline to least reduction in infections with respect to Baseline. The particles themselves are ordered on the vertical axis by their ranking for the Central 60% coverage, LAIV to 5–<18 scenario.

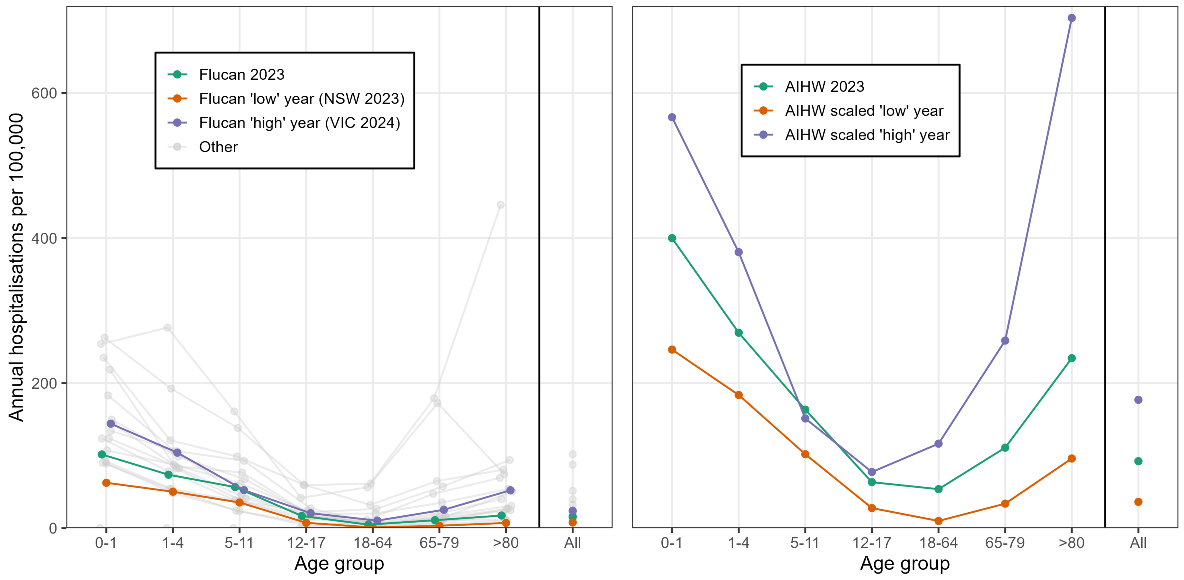


Figure S6: Hospitalisation rates per 100,000 of the population (in each age group) which were used to calculate infection hospitalisation rates for each model particle (see Methods). (Left) Hospitalisation rates per age group in FluCan data. Hospitalisation rates are shown for each jurisdiction (eight) and year (2023 or 2024) combination (grey). Hospitalisation rates at the national level are shown for 2023 (green, used to compare FluCan with AIHW data), and the jurisdiction and year combinations selected as a 'high' (green) and 'low' (orange) hospitalisation burden year. (Right) Hospitalisation rates per age group in AIHW data for 2023 (green), along with hospitalisation rates used in sensitivity analysis to represent 'high' and 'low' burden years; these were calculated by scaling the AIHW 2023 rates by age-specific multiplicative factors based on the FluCan data (see Methods).

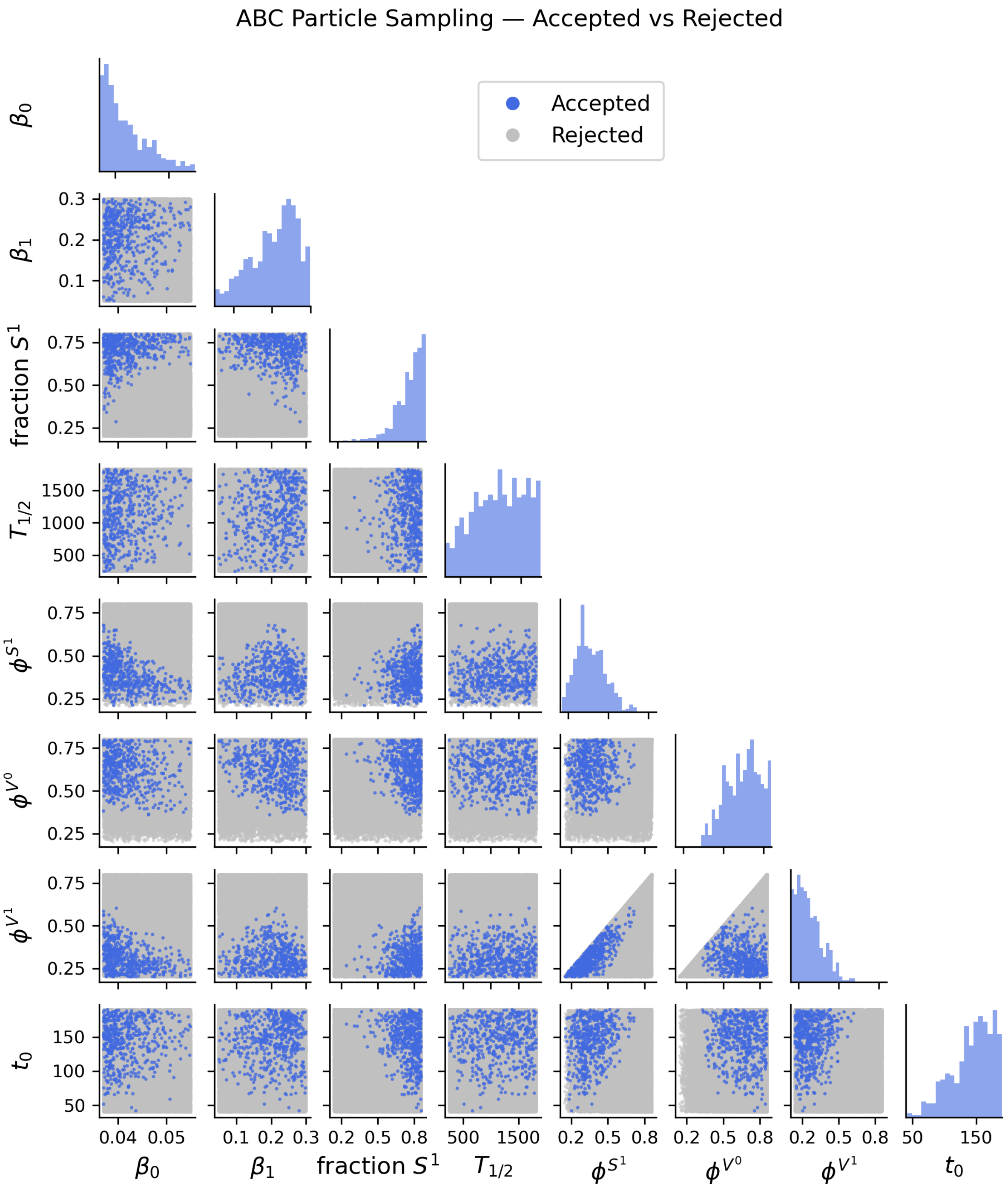


Figure S7: Correlation plot of parameters calibrated via ABC. These are the seasonality parameters ($\beta_0$, $\beta_1$, and $t_0$), the initial fraction of the population with pre-existing immunity, the half-life of the the waning of the pre-existing immunity ($T_{1/2}$), and the susceptibility to infection reductions of the compartments with pre-existing immunity ($\varphi^{S^1}$), vaccination ($\varphi^{V^0}$), or both ($\varphi^{V^1}$).

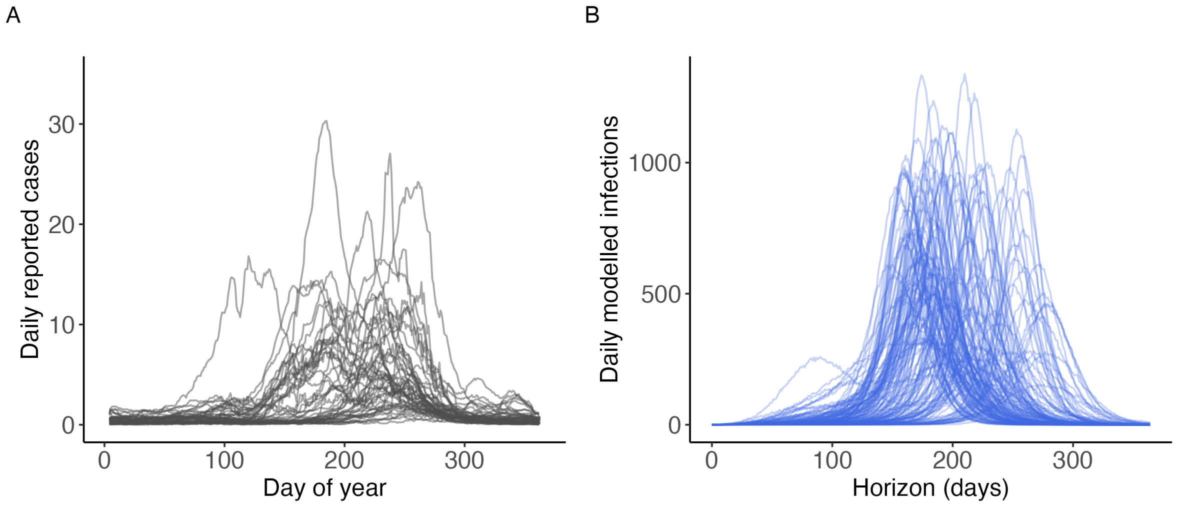


Figure S8: **A** Data on daily reported cases of influenza per 100,000 population. Each line represents a single combination of year (from 2015–2025, excluding 2020–2023) and Australian state/territory (all states/territories except the Northern Territory, which does not exhibit seasonality in influenza case patterns). Case data were sourced from the Australian National Notifiable Diseases Surveillance System (NNDSS). **B** Simulated daily infection incidence.

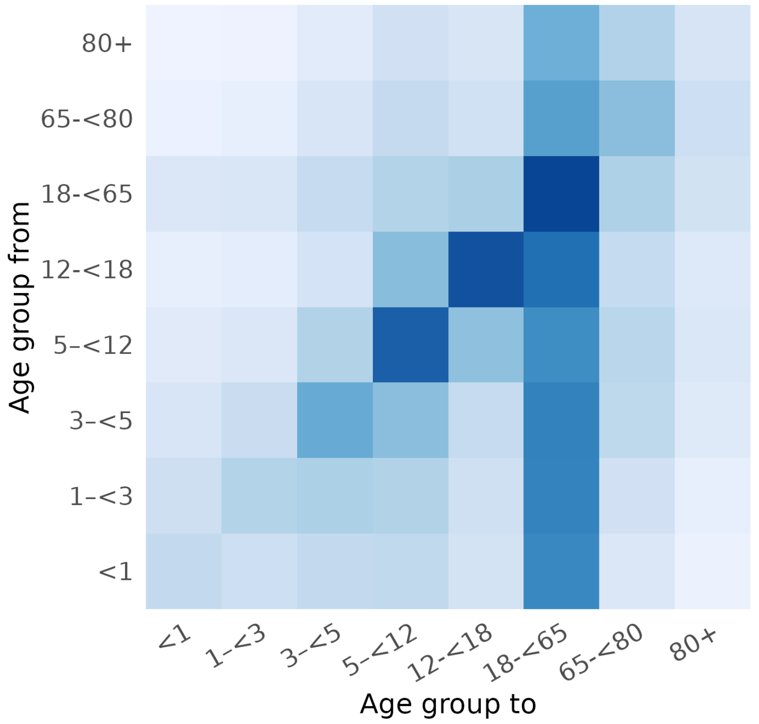


Figure S9: Age-specific contact matrix used in the transmission model, derived from the POLYMOD study and aggregated to the eight age groups considered in this work.

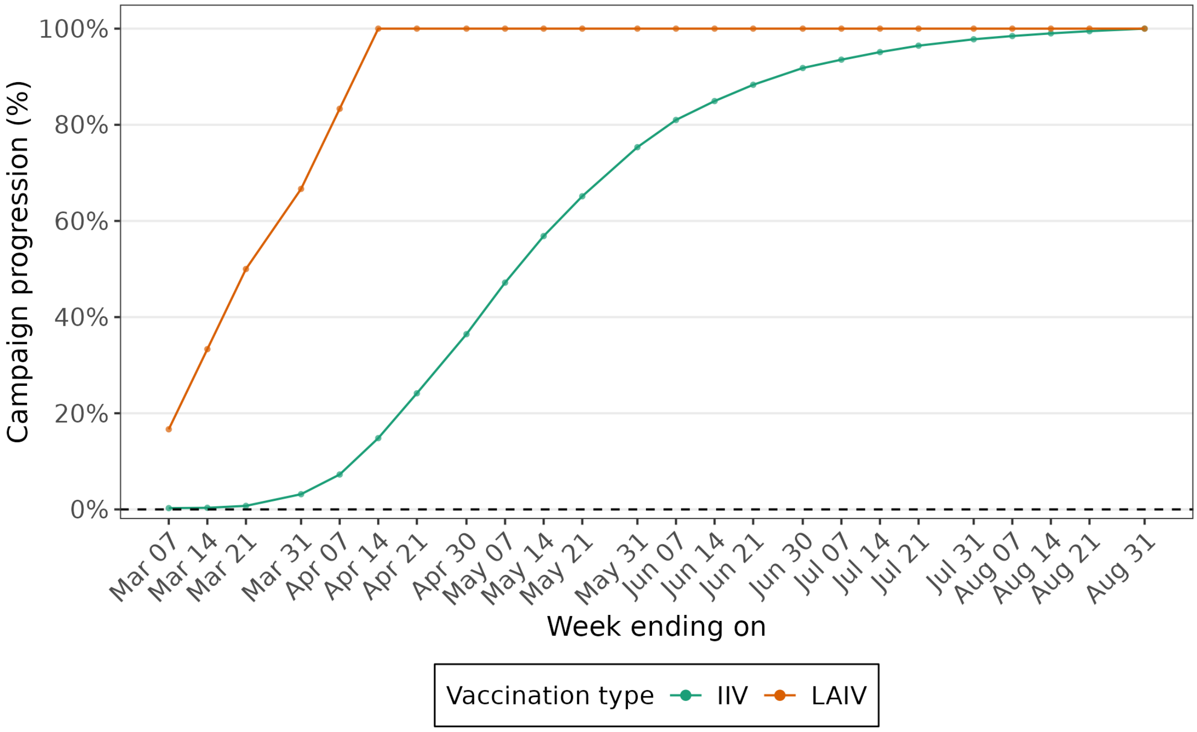


Figure S10: Progression of the modelled vaccination campaigns over time. Two vaccines are delivered: IIV over 24 weeks, and LAIV over 6 weeks (to primary and secondary school children).

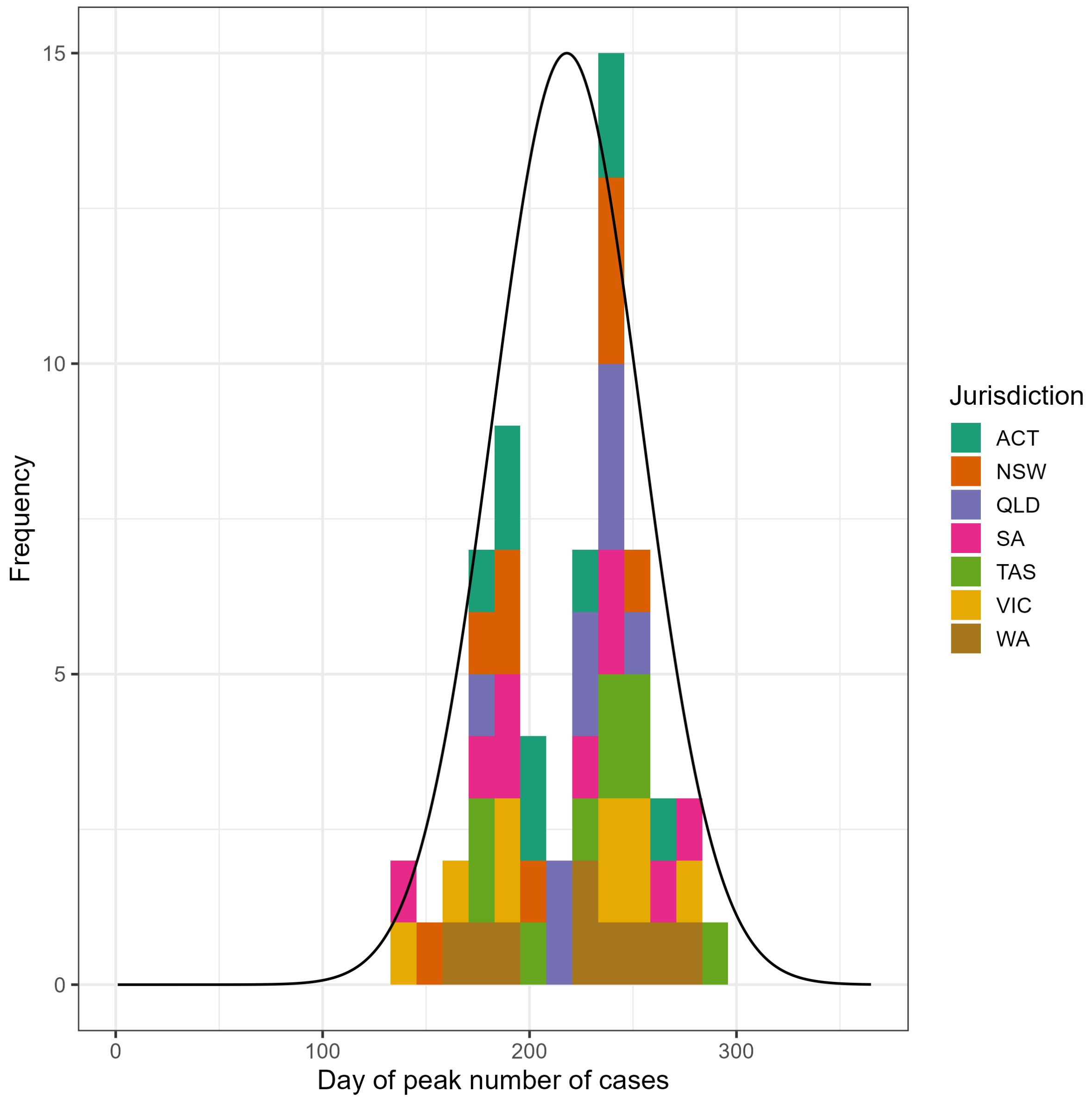


Figure S11: Distribution of the timing of the seasonal influenza peak in Australian jurisdictions. Histogram shows the day of the year of peak case numbers by jurisdiction, using 5-day bins. Data from the 2015–2019 and 2022–2025 influenza seasons were included, excluding the period during which COVID-19 restrictions substantially disrupted influenza transmission. All Australian states and territories were included except the Northern Territory, whose influenza dynamics are not characterised by strong seasonal forcing. For each season and jurisdiction, we identified the day of maximum daily influenza case incidence. The solid line shows a normal distribution with mean day 218 and standard deviation 36 days, rescaled vertically for visual comparison. The ABC acceptance range for peak timing was defined as the mean $\pm 1.96$ standard deviations.

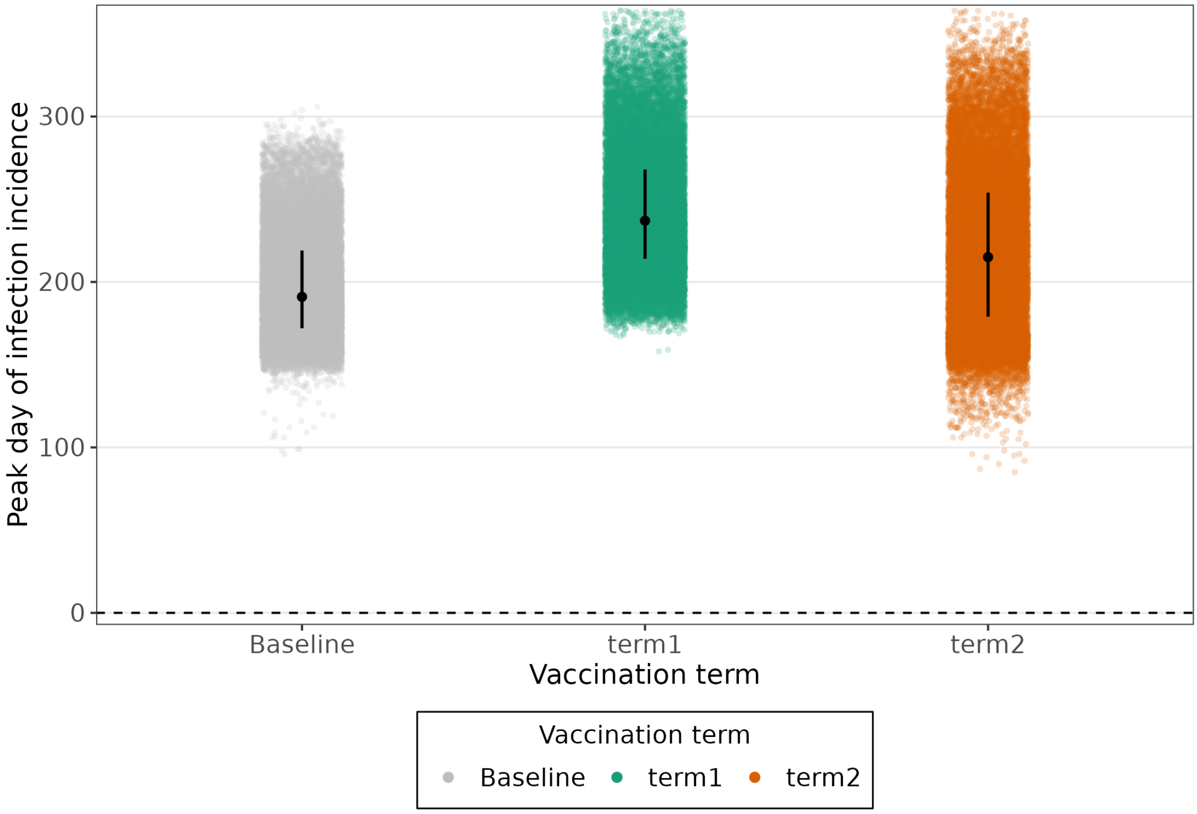


Figure S12: Day of peak of infection incidence for Baseline (i.e. no LAIV), term 1 vaccination (days 59–100), and term 2 vaccination (days 110–170). For some model particles, infections were already increasing when term 2 vaccination started. It therefore had less impact on early transmissions and consequently, epidemics were not shifted as late in the season, as can be seen by the median peak day and its IQR (black dot and vertical lines).

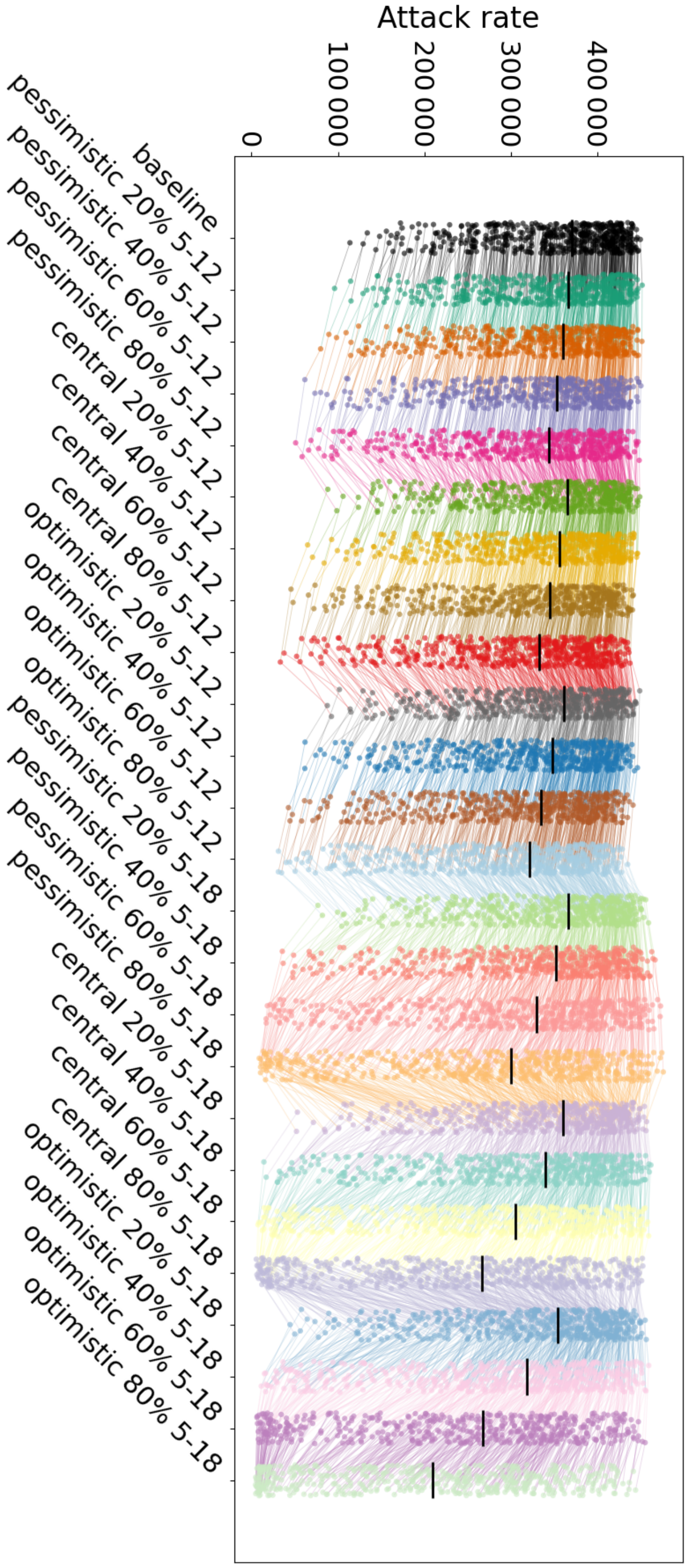


Figure S13: Infection attack rates across all LAIV intervention scenarios. Coloured points show the mean infection attack rate for each model particle across its 50 stochastic simulation runs, with lines connecting results for the same particle across scenarios. Black lines indicate the median across all particles for each scenario.

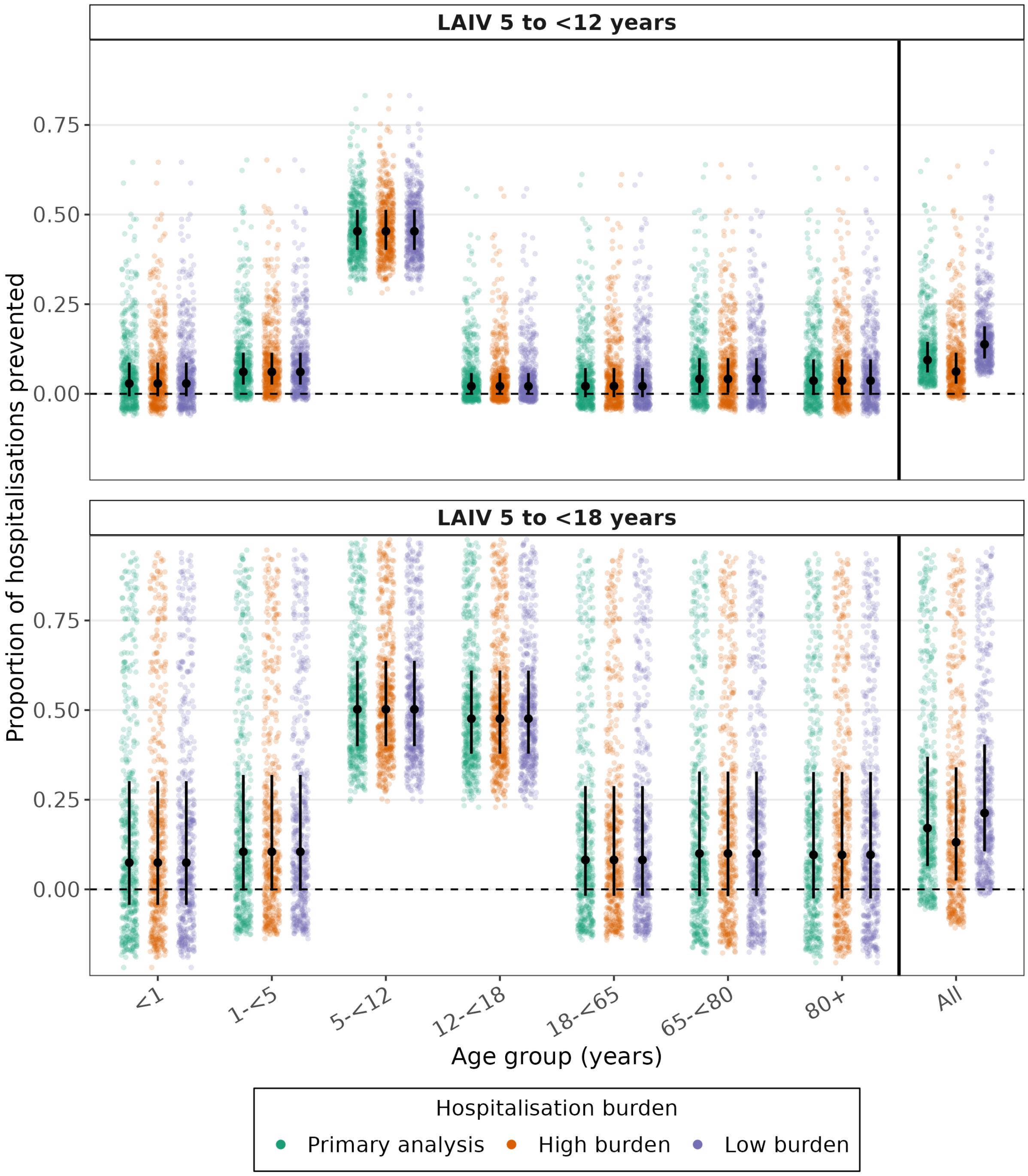


Figure S14: Estimated proportion of in influenza-associated hospitalisations prevented under school-based LAIV rollout (Central VE, 60% LAIV coverage), by age group and hospitalisation burden, relative to Baseline scenario for LAIV rollout targeting children aged 5–<12 years (top) and 5–<18 years (bottom). Coloured points show median outcomes from all runs for each particle, stratified by age group. Black points and vertical lines show the median and IQR, respectively. Estimates are shown using hospitalisation rates derived from AIHW 2023 data (green), a 'high' burden season (FluCAN data adjusted for 2024 in the state of Victoria) (orange), and a 'low' burden season (FluCAN data adjusted for 2023 in the state of New South Wales) (purple), by age group and for the population overall. The horizontal dashed line indicates no change in hospitalisations relative to the status quo. Negative values indicate an increase in hospitalisations.

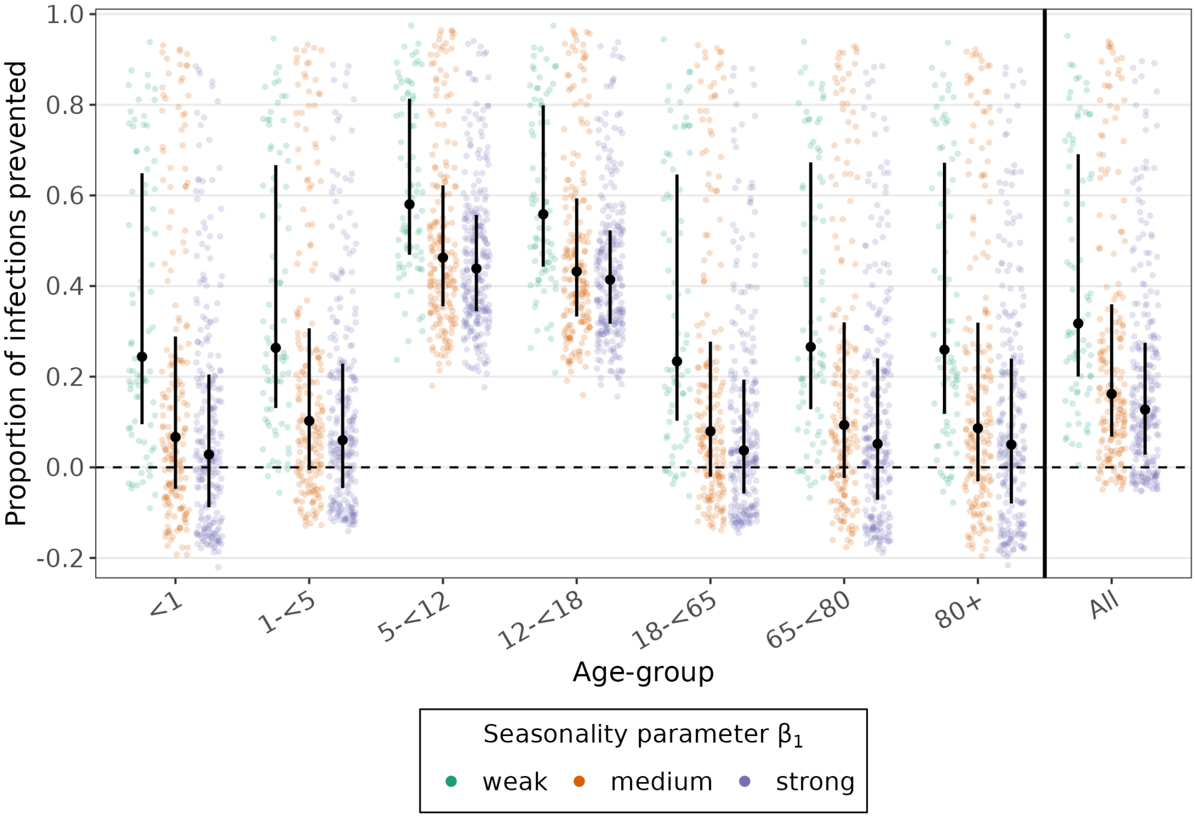


Figure S15: Sensitivity analysis of the proportion of infections prevented (compared to Baseline) in the scenario with Central VE and 60% LAIV coverage in primary and secondary school children. Coloured points show median outcomes from all runs for each particle, classified according to the seasonality parameter $\beta_1$. Black points and vertical lines show the median and IQR, respectively. Accepted $\beta_1$ values range from 0.05 to 0.30 and are divided into three equal-width intervals, labelled ”weak”, ”medium”, and ”strong”.

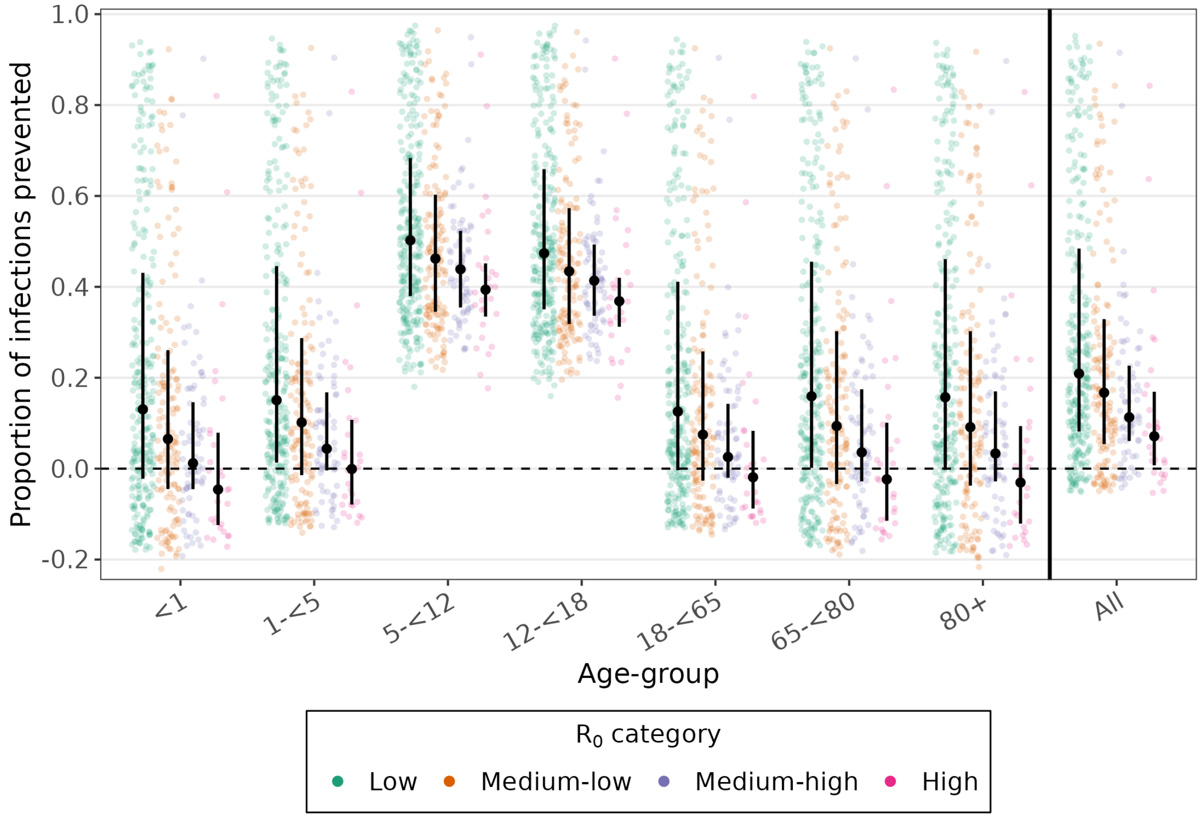


Figure S16: Sensitivity analysis of the proportion of infections prevented (compared to Baseline) in the scenario with Central VE and 60% LAIV coverage in primary and secondary school children. Coloured points show median outcomes from all runs for each particle, classified according to $R_0$ calculated at **mean transmissibility**. Black points and vertical lines show the median and IQR, respectively. We divided the range of this $R_0$ (1.57–2.34) into four bins: $R_0 < 1.75$ (Low), $1.75 \leq R_0 < 1.95$ (Medium-low), $1.95 \leq R_0 < 2.15$ (Medium-high), $2.15 \leq R_0$ (High).

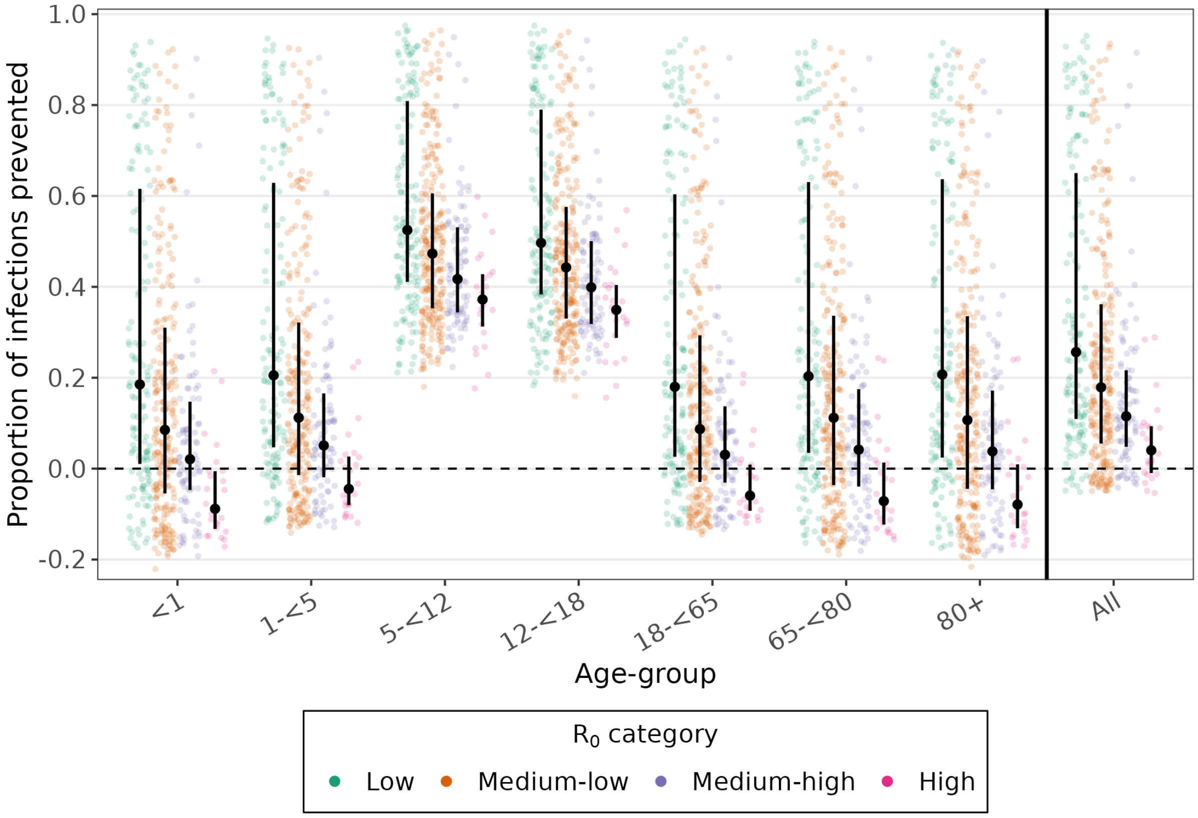


Figure S17: Sensitivity analysis of the proportion of infections prevented (compared to Baseline) in the scenario with Central VE and 60% LAIV coverage in primary and secondary school children. Coloured points show median outcomes from all runs for each particle, classified according $R_0$ calculated at **maximal transmissibility**. Black points and vertical lines show the median and IQR, respectively. We divided the range of $R_0$ calculated at **maximal transmissibility** (1.67–3.00) into four bins: $R_0 < 2.00$ (Low), $2.00 \leq R_0 < 2.33$ (Medium-low), $2.33 \leq R_0 < 2.66$ (Medium-high), $2.66 \leq R_0$ (High).

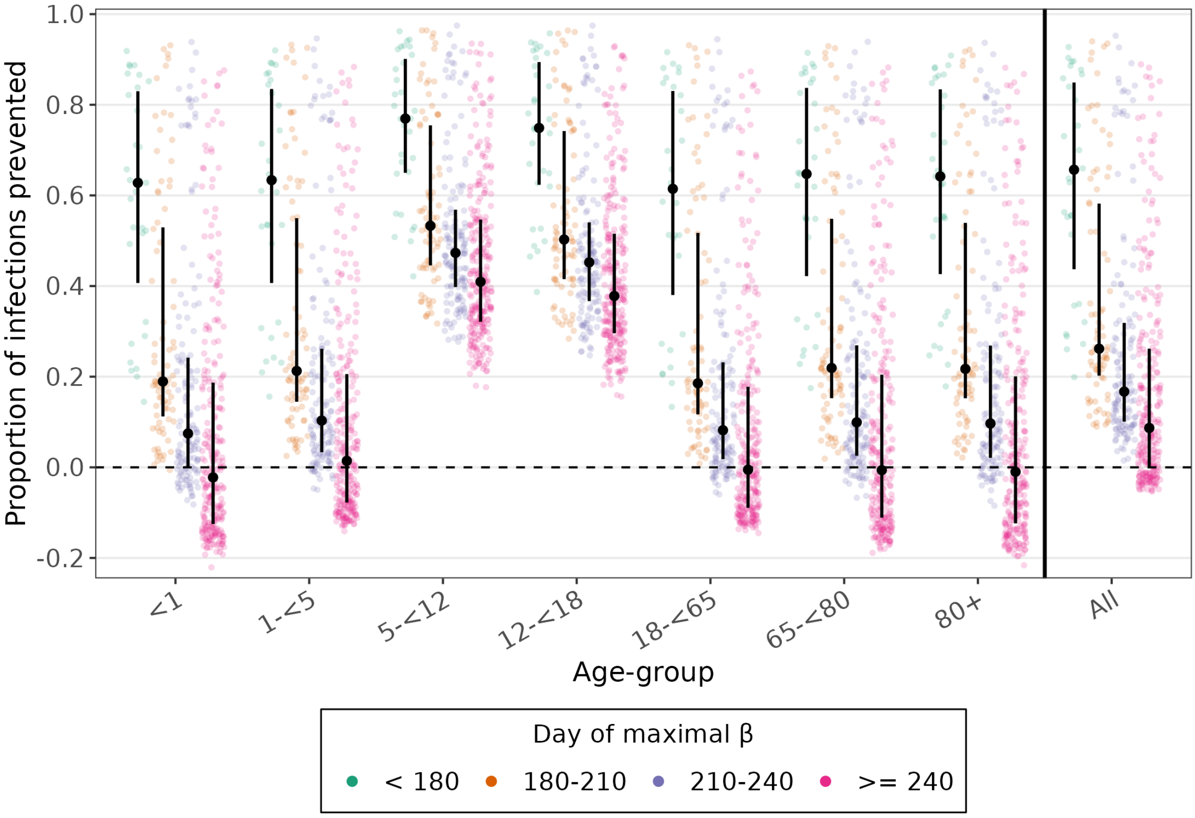


Figure S18: Sensitivity analysis of the proportion of infections prevented (compared to Baseline) in the scenario with Central VE and 60% LAIV coverage in primary and secondary school children. Coloured points show median outcomes from all runs for each particle, classified by the day at which the transmissibility $\beta$ reaches its maximum. Black points and vertical lines show the median and IQR, respectively.More infections are prevented across all age groups for epidemics for which the transmissibility peaks earlier on in the season.

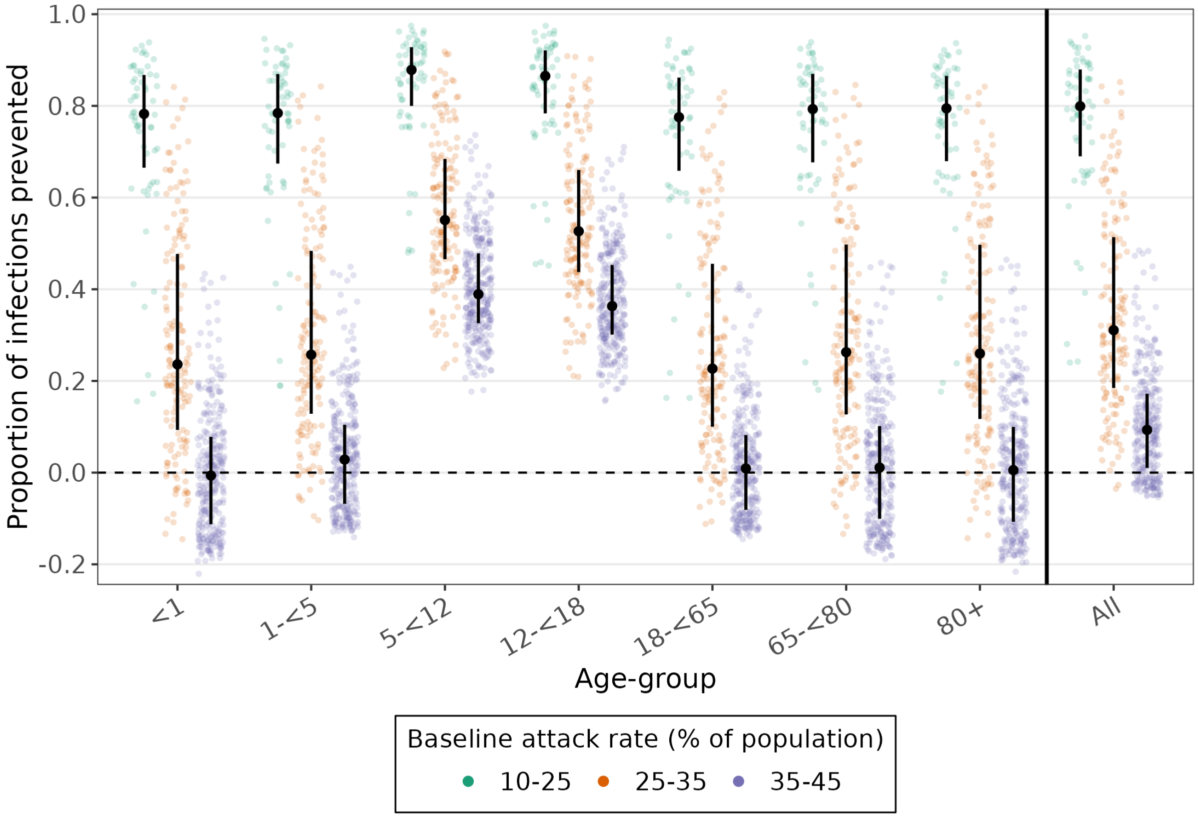


Figure S19: Sensitivity analysis of the proportion of infections prevented (compared to Baseline) in the scenario with Central VE and 60% LAIV coverage in primary and secondary school children. Coloured points show median outcomes from all runs for each particle, classified according to the Baseline attack rate (as a % of the population). Black points and vertical lines show the median and IQR, respectively. Proportionally, more infections are prevented across all age groups for epidemics for which the Baseline infection attack rate is lower.

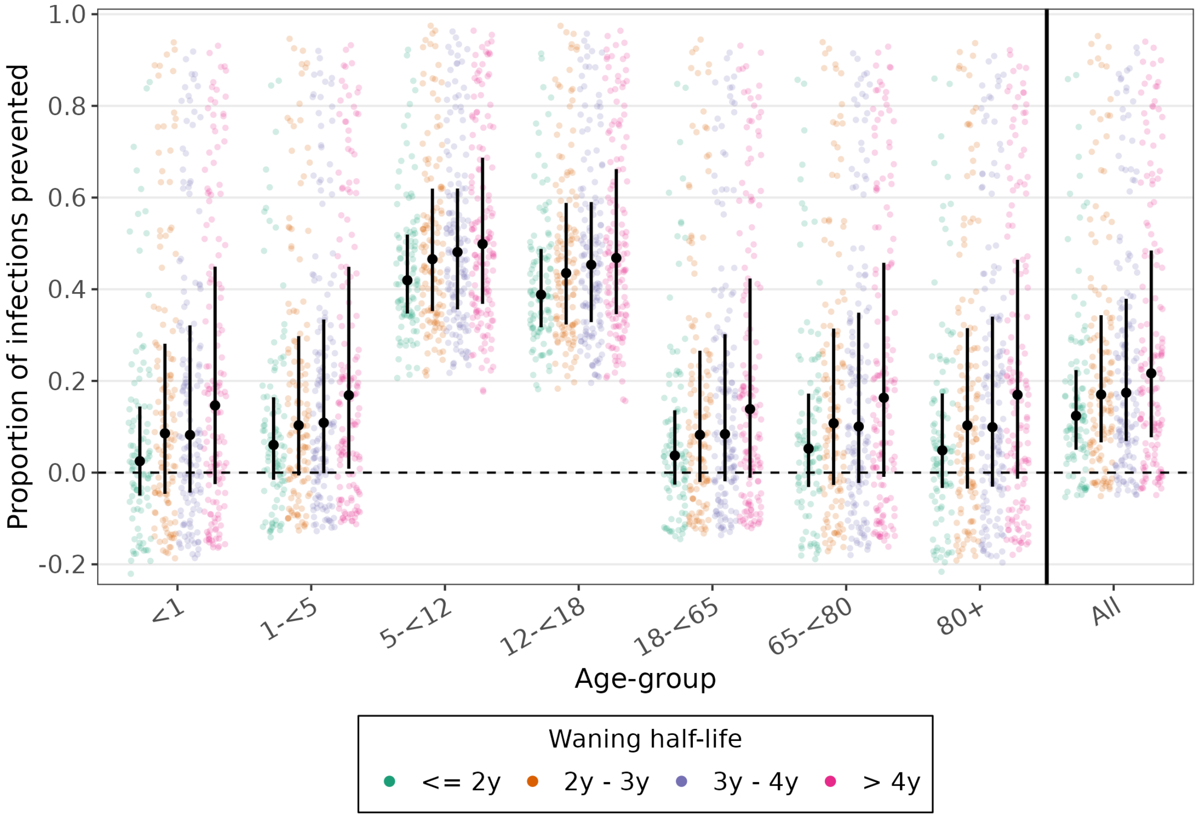


Figure S20: Sensitivity analysis of the proportion of infections prevented (compared to Baseline) in the scenario with Central VE and 60% LAIV coverage in primary and secondary school children. Coloured points show median outcomes from all runs for each particle, classified according to waning half-life of the pre-existing immunity. Black points and vertical lines show the median and IQR, respectively. When waning occurs slower (i.e. longer half-lives), a larger proportion of infections is prevented.

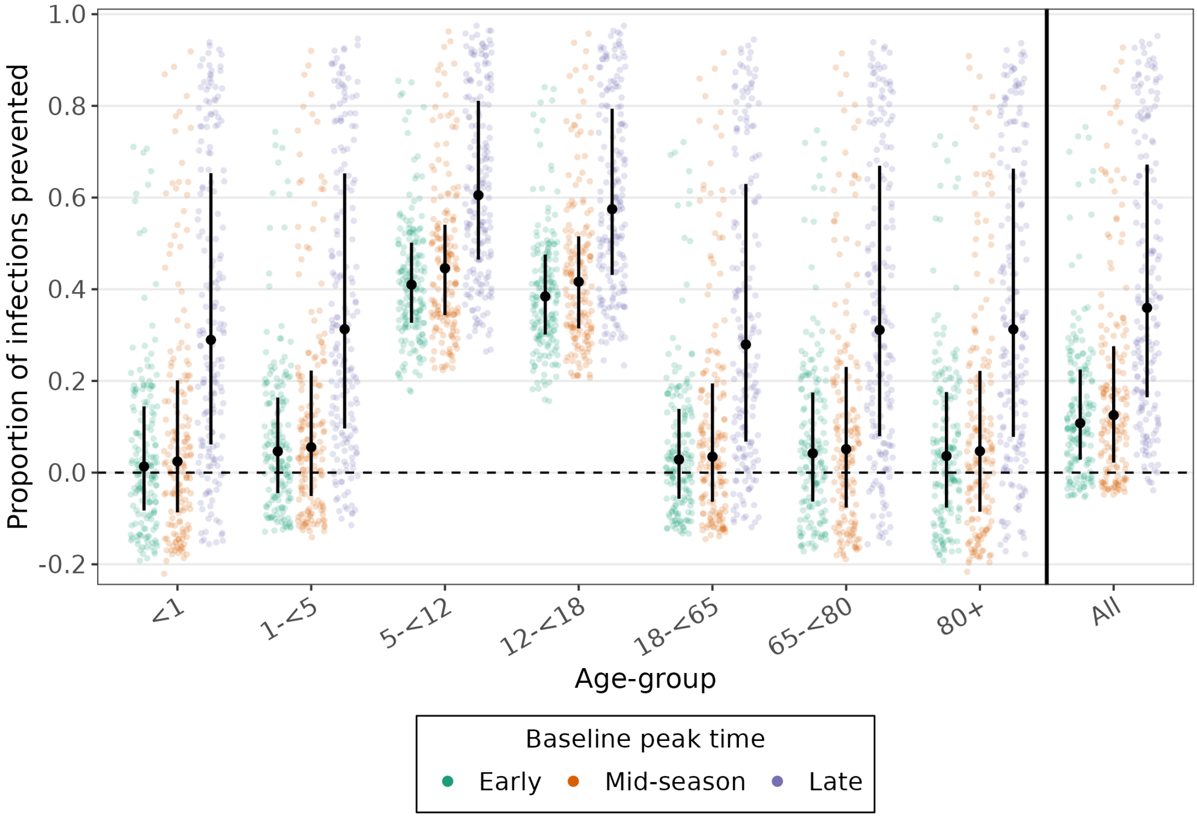


Figure S21: Sensitivity analysis of the proportion of infections prevented (compared to Baseline) in the scenario with Central VE and 60% LAIV coverage in primary and secondary school children. Coloured points show median outcomes from all runs for each particle, classified according to the Baseline peak time of infection incidence. Black points and vertical lines show the median and IQR, respectively. These median peak days ranged from day 149 to day 283. Particles were divided into three equally sized groups based on this value: Early (149–<178), Mid-season (178–<206), and Late (206–283). Particles in the Late group showed both a higher proportion of infections prevented and greater uncertainty in this proportion than particles in the Early and Mid-season groups.

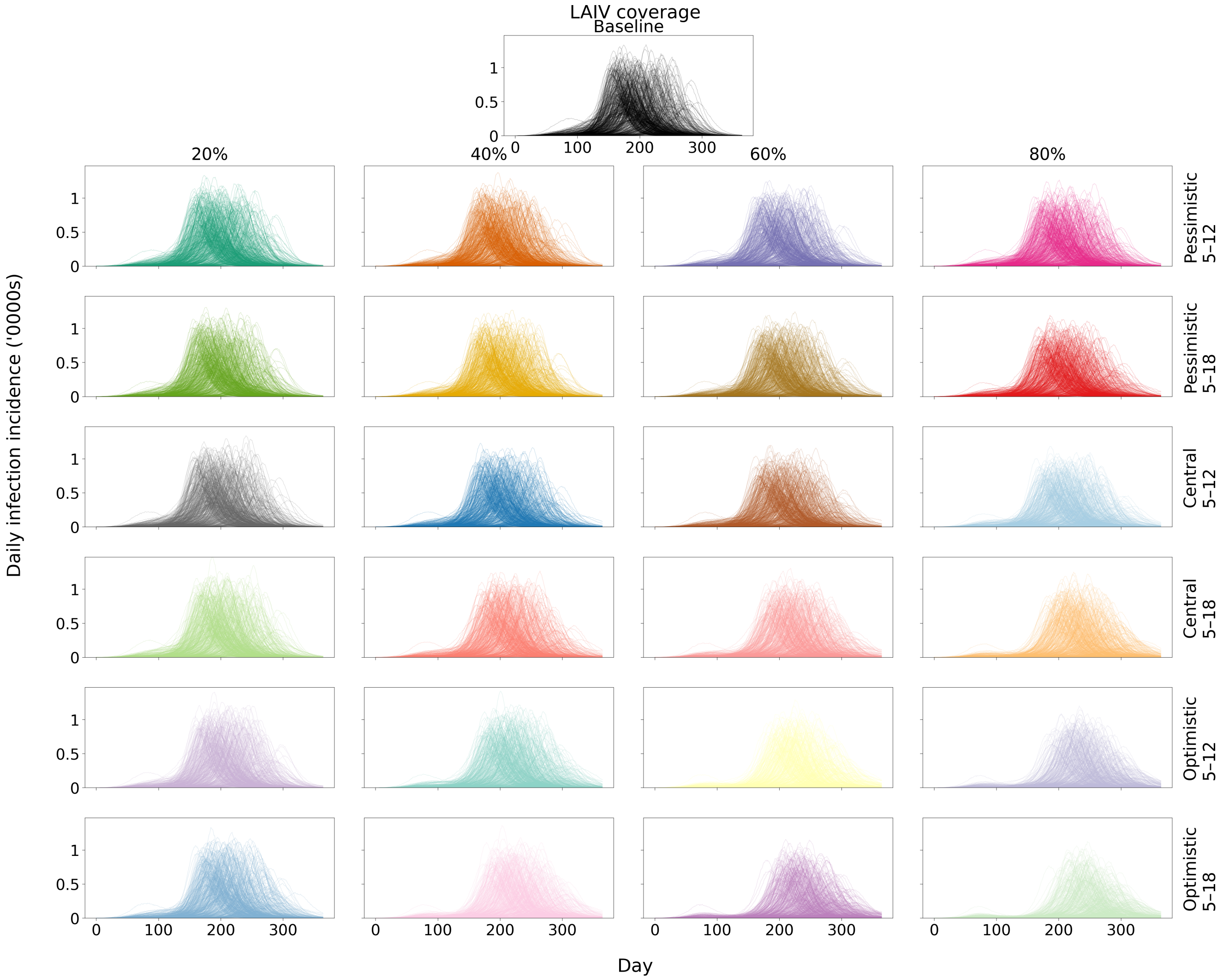


Figure S22: Daily influenza infection incidence across all LAIV scenarios. Each panel shows simulated daily infection incidence trajectories for individual accepted particles (one randomly selected run per particle). The Baseline scenario is shown at the top. Intervention scenarios vary by LAIV coverage (20%, 40%, 60%, and 80%), VE assumptions (Pessimistic, Central, and Optimistic), and target age group (5–<12 and 5–<18 years). Infection incidence is reported in thousands of infections.

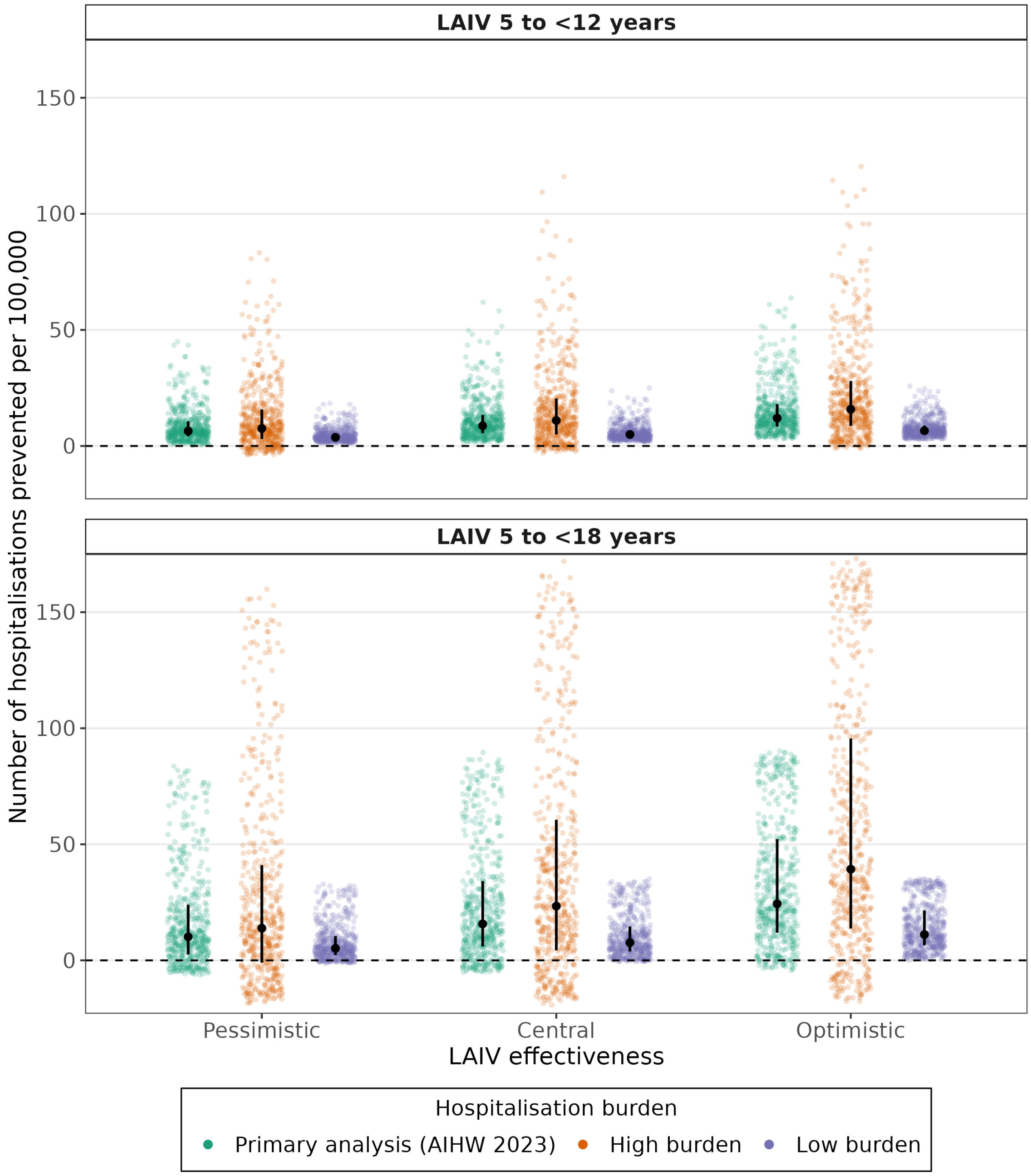


Figure S23: Estimated number of population-level influenza-associated hospitalisations prevented per 100,000 under school-based LAIV rollout (Central VE, 60% LAIV coverage), by LAIV VE assumptions and hospitalisation burden, relative to Baseline scenario for LAIV rollout targeting children aged 5–<12 years (top) and 5–<18 years (bottom). Coloured points show median outcomes from all runs for each particle. Black points and vertical lines show the median and IQR, respectively. Estimates are shown using hospitalisation rates derived from AIHW 2023 data (green), a 'high' burden season (FluCAN data adjusted for 2024 in the state of Victoria) (orange), and a 'low' burden season (FluCAN data adjusted for 2023 in the state of New South Wales) (purple), by age group and for the population overall. The horizontal dashed line indicates no change in hospitalisations relative to the status quo. Negative values indicate an increase in hospitalisations.

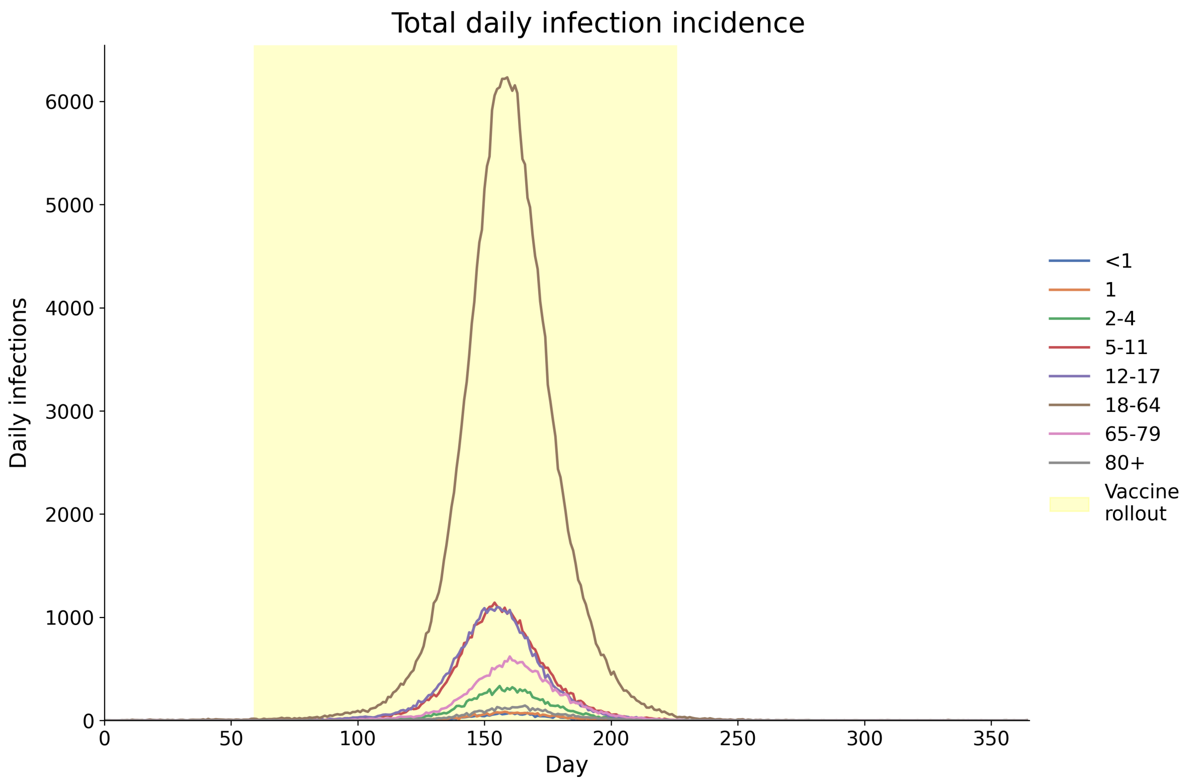


Figure S24: Daily infection incidence for each age group for a randomly selected accepted particle in the Baseline scenario.

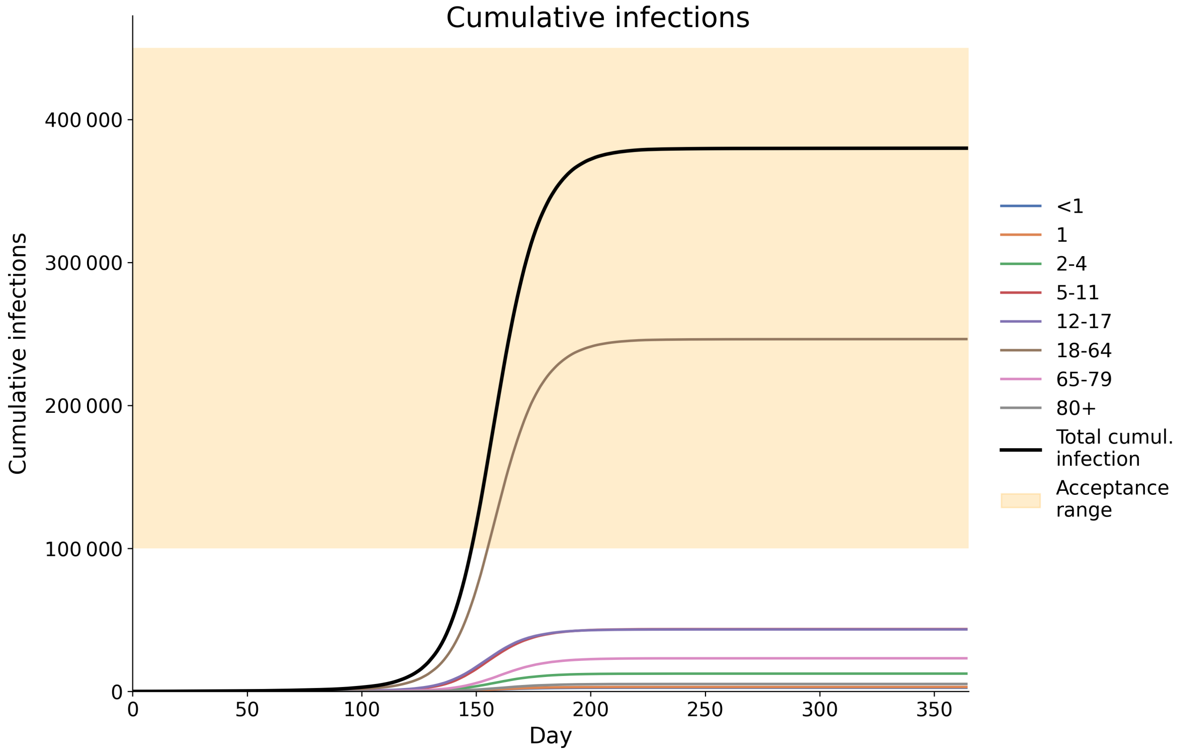


Figure S25: Cumulative infection incidence for each age group for a randomly selected accepted particle in the Baseline scenario (same particle as in Figure S24).

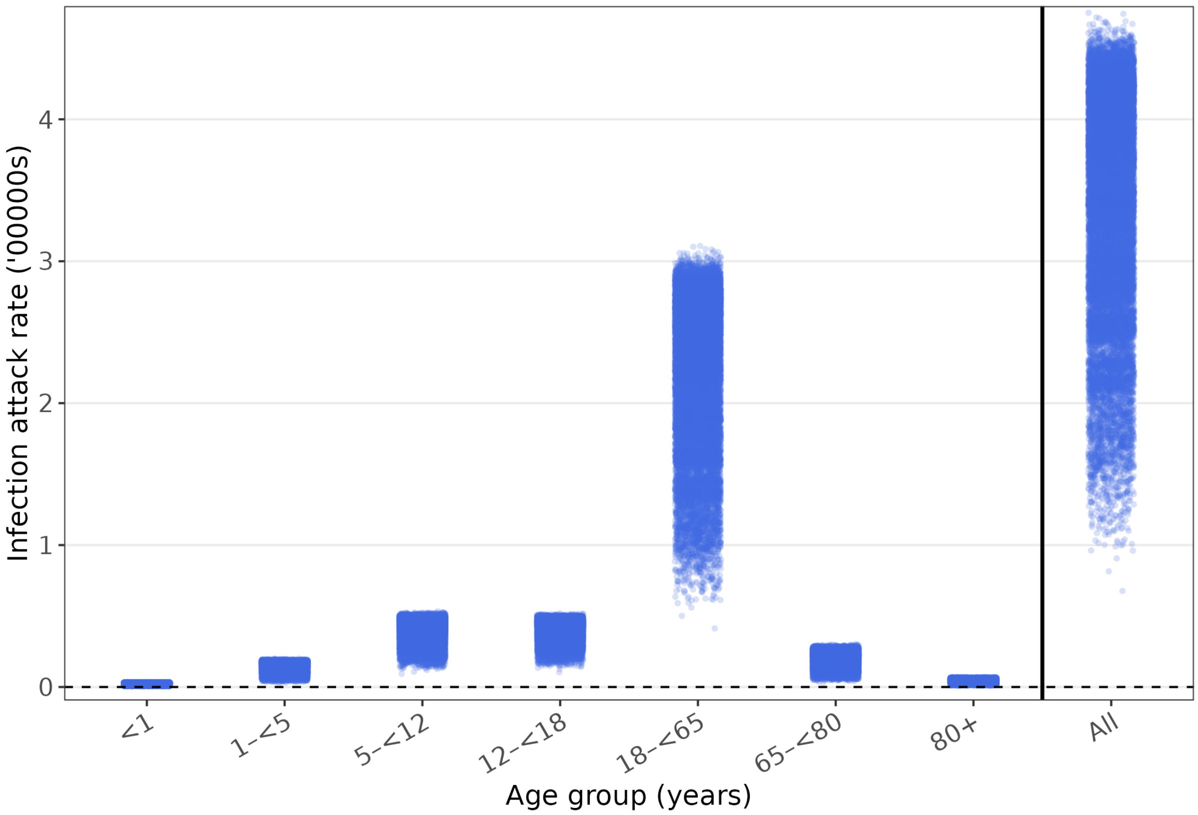


Figure S26: Infection attack rate for all Baseline runs (all 50 runs for all 518 accepted particles), stratified by age.

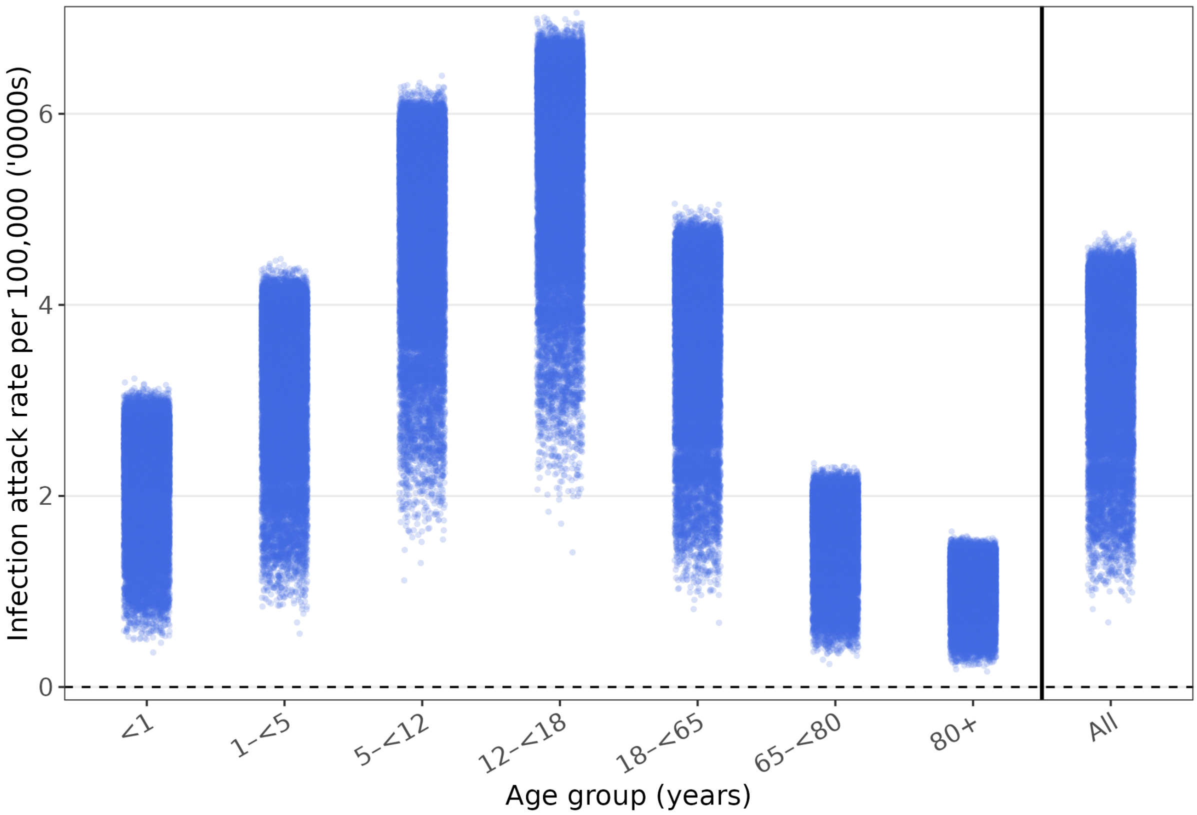


Figure S27: Infection attack (per 100,000) rate for all Baseline runs (all 50 runs for all 518 accepted particles), stratified by age.

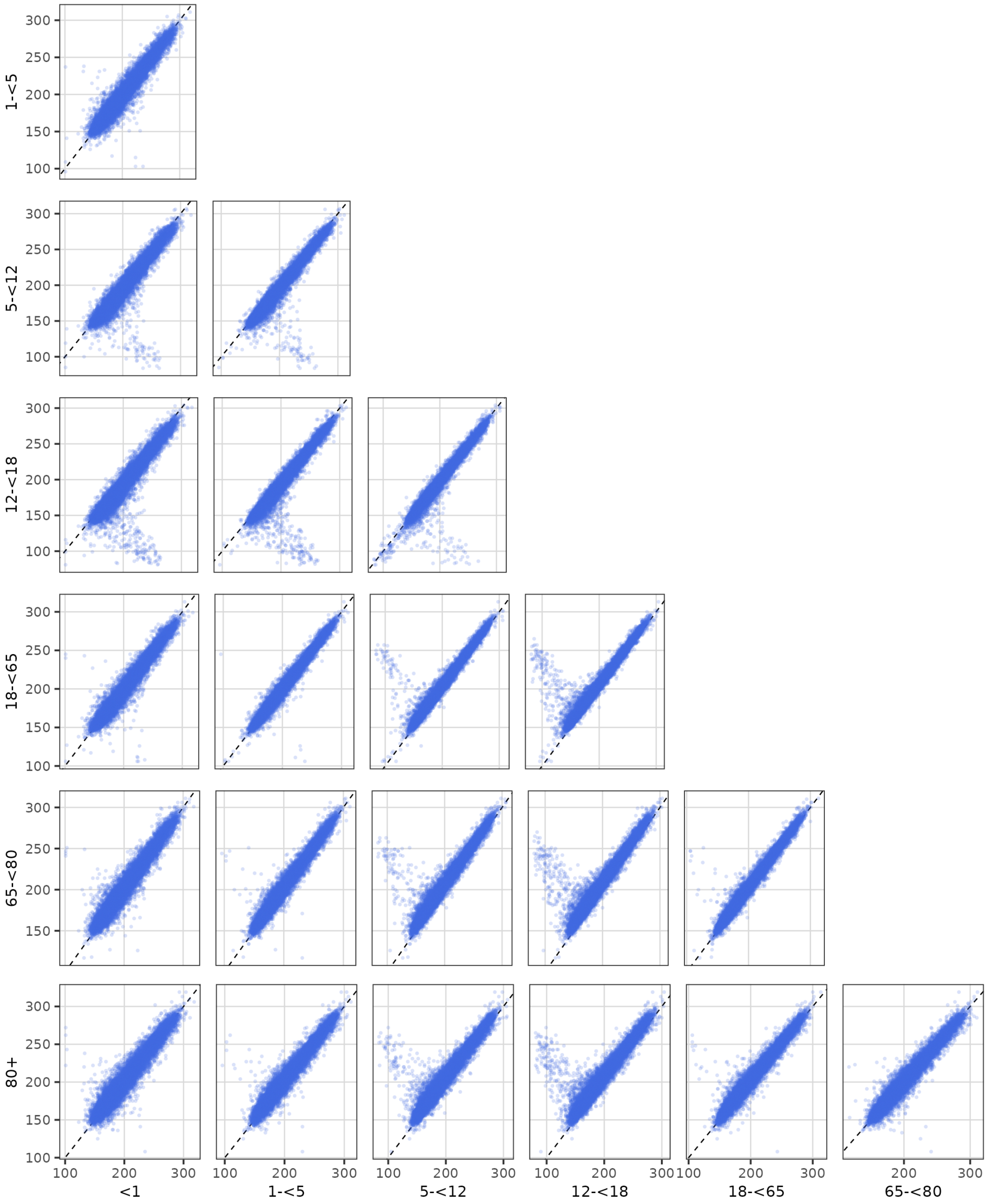


Figure S28: The day of peak infection incidence is shown for all Baseline runs, for each combination of age groups. Dots sitting below the dashed diagonal line are runs where the peak day of the infection incidence for the younger of the age groups occurred earlier.

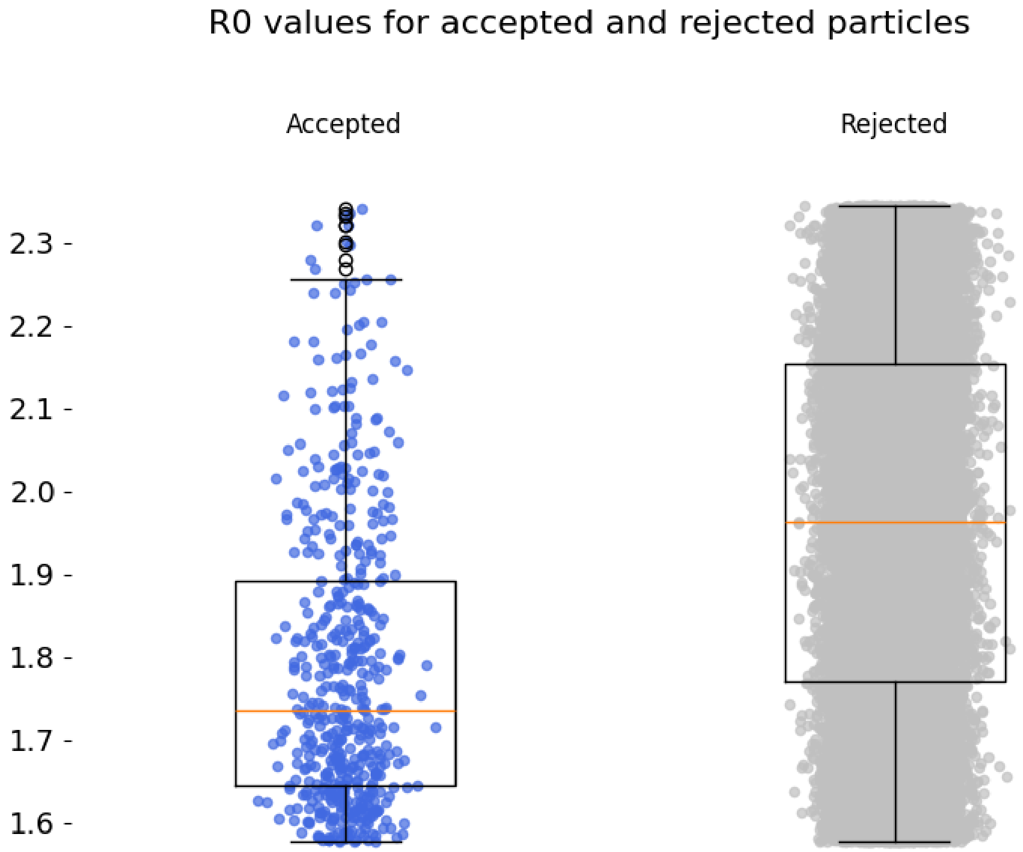


Figure S29: Values of $R_0$ corresponding to all 70,000 particles evaluated during the ABC step (for $K = 0$, i.e. the sine wave seasonal forcing), shown separately for the 518 accepted and 69,482 rejected particles.

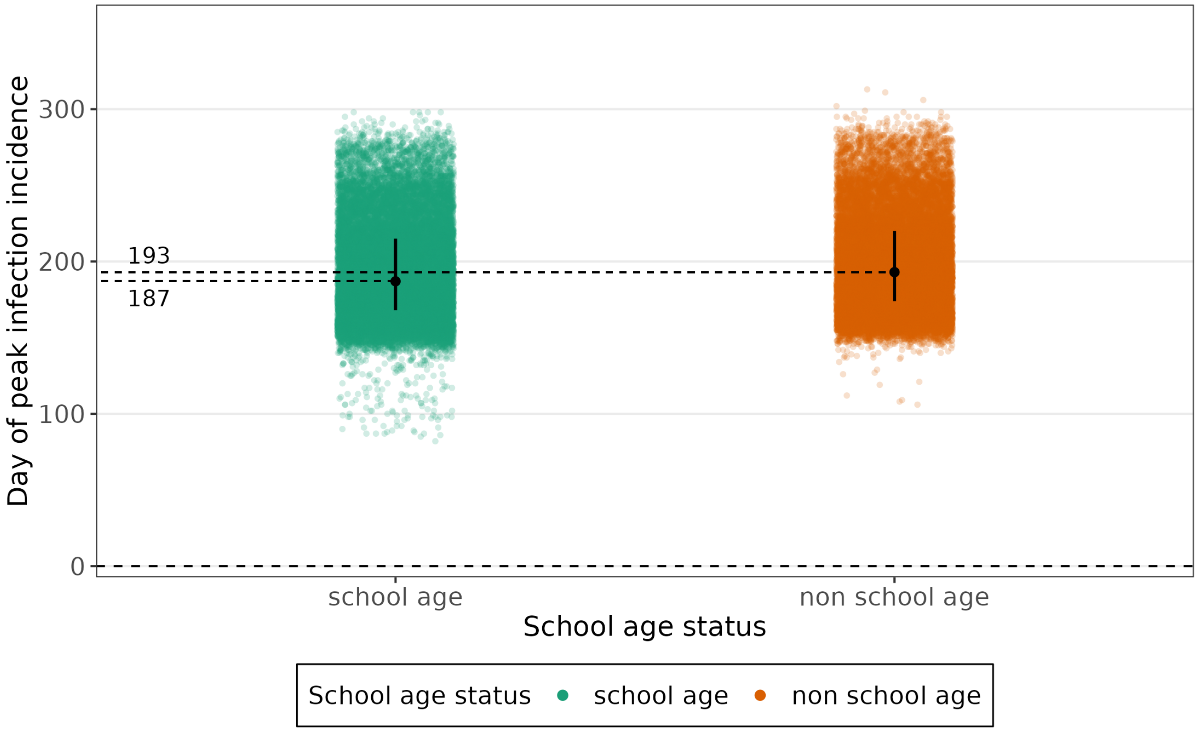


Figure S30: Comparison of peak day of infection incidence, stratified by school age (5–<18) vs. non school age (0–<5 and 18+). The median peak day for school age children is approximately 1 week earlier than for the rest of the population.

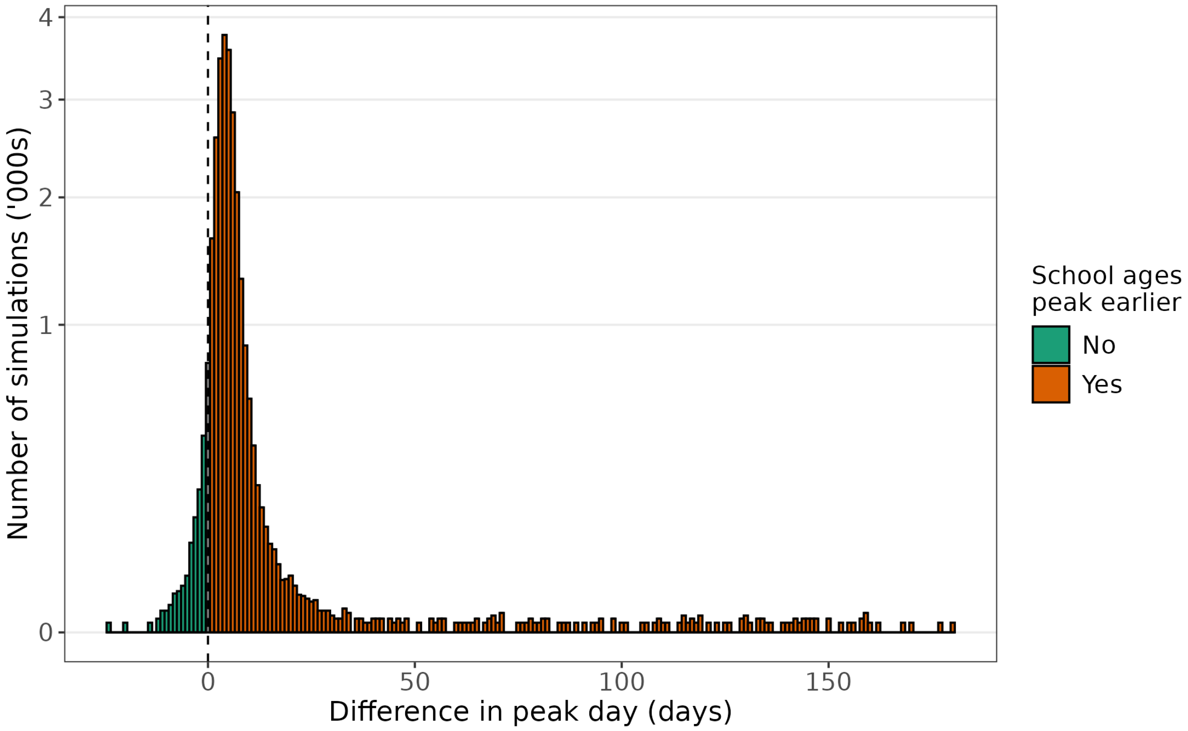


Figure S31: Comparison of peak infection incidence between school age (5–$¡$18 years) and non school age (0–<5 and 18+ years) groups. The histogram shows the difference in peak timing, expressed as the number of days by which infection incidence peaked later in the non school age group than in the school age group. The median difference was 4 days (IQR: 3–6 days).

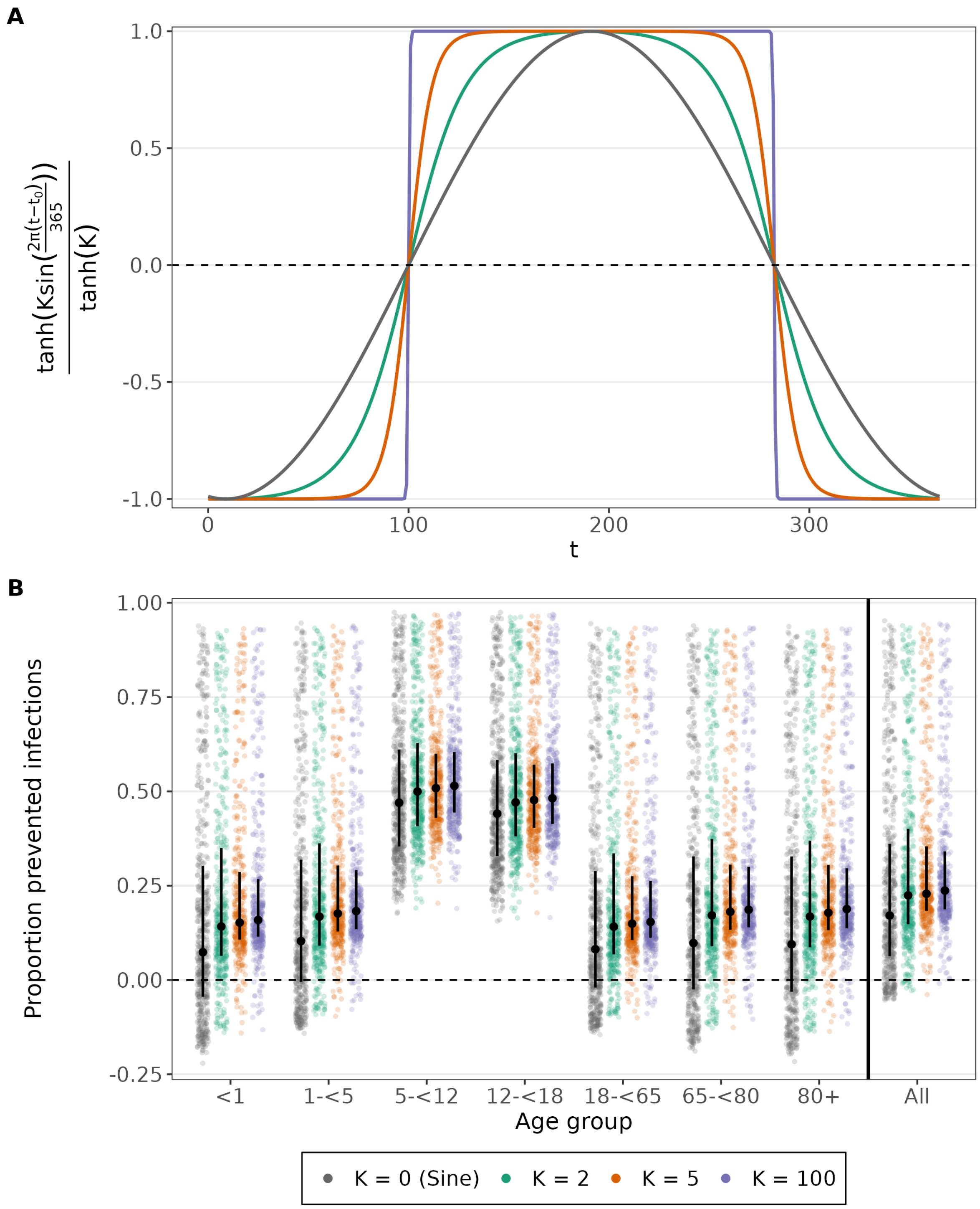


Figure S32: **A** Seasonal forcing $\beta_K(t) = \tanh(K\sin(\frac{2\pi(t-t_0)}{365}))/\tanh(K)$ for different values of K. The parameter K controls the shape of the seasonal forcing, interpolating between sinusoidal forcing as $K \to 0$ and square wave forcing as $K \to \infty$. **B** Age-stratified proportion of prevented infections for Central VE, cover 60 % administered to 5–<18, between sinusoidal wave forcing and square wave forcing. The median percentage of prevented infections for the whole population increases from 17.10% ($K = 0$) to 23.71% ($K = 100$).

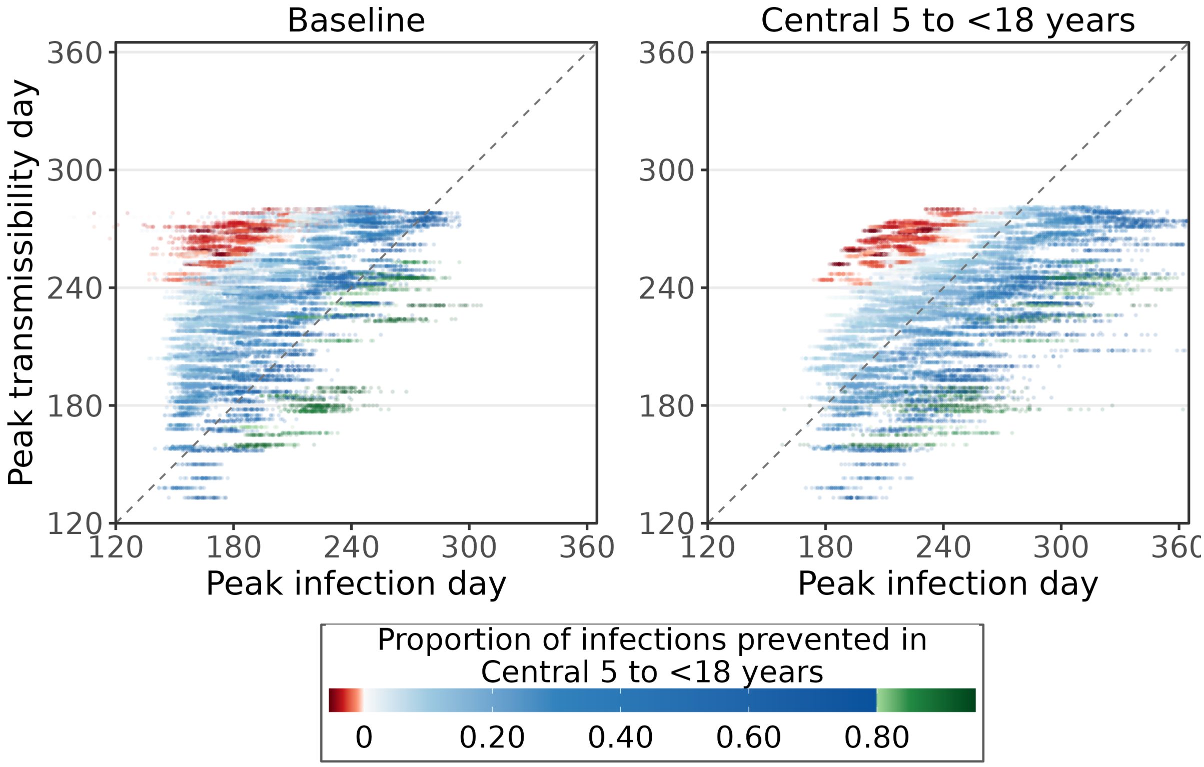


Figure S33: (Day of peak of transmissibility (i.e. peak of $\beta(t)$) versus day of peak infection incidence. Each of the 518 accepted particles is represented by 50 points, corresponding to its 50 simulation runs. Colour indicates the proportion of infections prevented in the 60% LAIV coverage, 5–<18, Central LAIV effectiveness scenario relative to Baseline. The runs with an increase in infections compared to Baseline are shown in red, while the ones with a decrease of at least 80% are shown in green. Particles associated with in increase in infections are clustered in the top left, corresponding to an early peak in infection incidence and late peak in transmissibility. While those associated with a strong suppression of of infections are clustered in the bottom right, corresponding to a late peak in infection incidence and early peak in transmissibility.
